# Omic Risk Scores are Associated with COPD-related Traits Across Three Cohorts

**DOI:** 10.1101/2025.06.01.25328699

**Authors:** Iain R. Konigsberg, Luciana B. Vargas, Katherine A. Pratte, Daniel E. Guzman, Tess D. Pottinger, Kristina L. Buschur, Thomas W. Blackwell, Yongmei Liu, Kent D. Taylor, W. Craig Johnson, Peter Durda, Russell P. Tracy, Ani Manichaikul, Elizabeth C. Oelsner, Stacey Gabriel, Namrata Gupta, Suna Onengut-Gumuscu, Joshua D. Smith, Francois Aguet, Kristin G. Ardlie, Usman A. Tahir, Robert E. Gerszten, Clary Clish, Eugene R. Bleecker, Deborah A. Meyers, Victor E. Ortega, Stephanie A. Christenson, Dawn L. DeMeo, Brian D. Hobbs, Craig P. Hersh, Peter J. Castaldi, Jeffrey L. Curtis, R. Graham Barr, Jerome I. Rotter, Stephen S. Rich, Prescott G. Woodruff, Edwin K. Silverman, Michael H. Cho, Katerina J. Kechris, Russell P. Bowler, Ethan M. Lange, Leslie A. Lange, Matthew R. Moll, NHLBI Trans-Omics for Precision Medicine (TOPMed) Consortium

## Abstract

**Background:** Chronic obstructive pulmonary disease (COPD) exhibits marked heterogeneity in lung function decline, mortality, exacerbations, and other disease-related outcomes. Omic risk scores (ORS) estimate the cumulative contribution of omics, such as the transcriptome, proteome, and metabolome, to a particular trait. This study evaluates the predictive value of ORS for COPD-related traits in both smoking-enriched and general population cohorts.

**Methods:** ORS were developed and tested in 3,339 participants of Genetic Epidemiology of COPD (COPDGene) with blood RNA-sequencing, proteomic, and metabolomic. Single- and multi-omic risk scores were trained 24 cross-sectional and five longitudinal traits using 80% of the data, focusing on disease severity, exacerbations, and traits from spirometry and computed tomography scans. Multivariable models were used to test ORS associations with outcomes in remaining COPDGene participants and externally validated in SubPopulations and InteRmediate Outcome Measures in COPD Study (SPIROMICS) (n = 2,177) and Multi-Ethnic Study of Atherosclerosis (MESA) (n = 1,000).

**Results:** In the COPDGene testing set, 69 of 72 single-omic ORS showed significant associations with 24 cross-sectional traits (adjusted *p*-value < 0·05). One of 15 longitudinal ORS was associated with changes in trait values between COPDGene visits. Significant associations were observed for all 38 cross-sectional ORS tested in SPIROMICS and for 16 of 24 in MESA. Proteomic and metabolomic risk scores generally displayed stronger associations than transcriptomic scores.

**Discussion:** Blood-based ORS can predict cross-sectional and future COPD-related traits in both smoking-enriched and general population cohorts.

## INTRODUCTION

Chronic obstructive pulmonary disease (COPD) is a progressive and debilitating respiratory condition that impacts the lives of millions of individuals worldwide.^1^ Characterized by persistent airflow limitation, COPD encompasses a group of chronic lung conditions, most notably chronic bronchitis and emphysema. While smoking is a major established risk factor, only a minority of smokers develop disease and there are no current curative pharmacological treatments.^2^ Given the marked heterogeneity in the presentation of COPD there is a need for improved methods to classify COPD patients.

Polygenic risk scores (PRS) have become a popular means of capturing complex disease risk.^3^ For most common, complex diseases, single polymorphisms are rarely useful in stratifying patients or serving as biomarkers, but in aggregate, in the form of PRS, are useful in stratifying patients by disease risk. We previously constructed a composite PRS of FEV_1_ and FEV_1_/FVC associated with COPD that has been replicated in multiple cohorts.^4^ This PRS has further been demonstrated to be associated with earlier COPD onset^5^ and to interact with pack-years of smoking on reduced lung function.^6^ However, although PRS have aided in disease classification, they generally explain limited additional trait variance in clinical risk models.

Studies like the Genetic Epidemiology of COPD (COPDGene) and SubPopulations and InteRmediate Outcome Measures in COPD Study (SPIROMICS)^7^ cohorts have generated high-throughput omics data on many participants. Analysis of these datasets has identified many candidate omics biomarkers for traits including COPD,^8^ emphysema,^9^ and FEV_1_ decline.^10^ Similarly to genetic variants, single blood biomarkers rarely capture an appreciable proportion of trait variance. Multiple studies have demonstrated that by aggregating multiple omic features into omic risk scores (ORS), these combined scores are much more predictive of traits than using a single or few analytes.^11^ By examining an individual’s unique molecular profile, ORS aim to provide a more accurate representation of disease susceptibility, progression, and response to treatment. Additionally, ORS can potentially be used as proxies for expensive or difficult-to-measure traits in additional cohorts. ORS have recently been developed for COPD-related traits. Promising early studies, including a transcriptomic risk score (TransRS) that predicted prospective lung function decline^12^ and metabolomic risk scores (MetRS) for lung function and lung density,^13^ indicate that ORS are useful in capturing COPD-related traits.

As more high-throughput blood-based omics become available in human disease cohorts, supported by initiatives like the Trans-Omics for Precision Medicine (TOPMed) program,^14^ it is possible to build and evaluate ORS using multiple omics technologies in thousands of individuals. In this work, we evaluated the utility of ORS to predict COPD-related traits by leveraging the detailed phenotyping from the COPDGene cohort (clinical, physiological, and imaging) to build and test a range of trait-specific MetRS, TransRS, proteomic risk scores (ProtRS), and multiomic risk scores (MultRS). We focused on COPD severity-related traits and traits derived from spirometry and high-resolution computed tomography (HRCT) images.^15, 16^ We replicated these ORS in two additional cohorts, SPIROMICS^7^ and the Multi-Ethnic Study of Atherosclerosis (MESA).^17^

## METHODS

### Cohorts

We used COPDGene as the discovery cohort for model development and evaluation. COPDGene is an ongoing multicenter COPD-enriched observational study of over 10,000 subjects that began enrollment in 2008. All participants had at least ten pack-years of cigarette smoking before enrollment; by design, there was a balance between self-identified non-Hispanic White (two-thirds) and African American (one-third) participants. COPDGene participants have completed three study visits (baseline, five-year follow-up, and ten-year follow-up). All omics data were generated from biospecimens obtained at the second study visit (five-year follow-up).

Replication was performed using omics data from the SPIROMICS and MESA cohorts. SPIROMICS is an ongoing multicenter observational study of COPD that enrolled 2,863 participants in 4 groups: severe COPD, mild/moderate COPD, smokers without airflow obstruction, and non-smoking controls. Participants underwent a baseline examination, followed by three annual follow-up visits and a fourth follow-up visit some time later. MESA is a population-based multicenter observational study that recruited participants who self-identified as White, African American, Hispanic, or Chinese American. Additional information on cohorts and omics processing is available in the **Supplementary Methods**.

### Cross-Sectional Measures

We trained multiple ORS on traits collected during the 2^nd^ COPDGene visit, with a focus on traits relating to COPD, including COPD case-control status (FEV_1_/FVC < 70%), spirometry measurements, and high-resolution computed tomography (HRCT) measurements. Additionally, we incorporated multiple measures of COPD severity/classification, including the Body mass index, airflow Obstruction, Dyspnea, and Exercise Capacity (BODE) index,^18^ St. George’s Respiratory Questionnaire (SGRQ) total score,^19^ and Global Initiative for Chronic Obstructive Lung Disease (GOLD) spirometry grades.^1^

HRCT traits consisted of airway wall thickness, volume-noise-bias corrected lung density,^20^ emphysema (defined as percentage of lung voxels with CT attenuation ≤-950 HU on inspiratory CT), functional residual capacity (FRC) estimated from expiratory CT, total lung capacity (TLC) estimated from inspiratory CT, FRC/TLC ratio, gas trapping (defined as percentage of lung voxels with CT attenuation ≤-856 HU on expiratory CT), square root of wall area of a hypothetical airway with internal perimeter of ten mm (Pi10), and wall area percent.

Spirometric traits were forced expiratory flow between 25% and 75% of vital capacity (FEF_25-75_), forced expiratory volume at one second (FEV_1_), forced vital capacity (FVC), FEV_1_/FVC, and peak expiratory flow (PEF). All spirometric traits were measured post-bronchodilator and analyzed as absolute values. The remaining traits consist of diffusing capacity of the lung for carbon monoxide (DLCO), presence of acute exacerbations, exacerbation frequency, resting oxygen saturation (SaO2), six-minute walk distance (6MWD), and presence of chronic bronchitis.

### Longitudinal Measures

We additionally prospectively assessed continuous longitudinal traits measuring change in trait value between five-year COPDGene visits (from Visit 2 to Visit 3 of the study). Traits included: 6MWD, lung density, FEV_1_ change (% predicted), FEV_1_ change (mL), and FEV_1_ change (mL/year).

### Study Design

COPDGene participants were randomly divided into training (80%) and testing (20%) sets. ORS were developed in the training set. Score performance was evaluated in the held-out COPDGene testing set, SPIROMICS, and MESA, where appropriate.

### Omics Input Data

Data from bulk RNA-sequencing, proteomic, and metabolomic analysis of peripheral blood was generated as described in **Supplementary Methods**.

### Score Generation

To develop ORS, we applied an elastic net regularized regression framework using the glmnet R package.^21, 22^ We fitted models to each trait for each omics dataset, providing a pool of all omics features as potential predictors. Continuous traits were natural-log-transformed if skewed. BODE index, SGRQ score, and exacerbation frequency were modeled using negative binomial distributions. We used ten-fold cross-validation to select models with the best-performing combination of hyperparameters. We identified the lowest mean-squared error (MSE) provided by a hyperparameter combination, and then selected the least complex model (maximum α and λ) within one standard error of best model MSE. This optimal model was then used to calculate ORS in the held-out COPDGene testing set and external replication cohorts.

### Score Evaluation

Scores were tested for association with traits of interest using generalized linear models with the trait as the outcome, ORS as a predictor, and adjusting for age, sex, race, smoking status, pack-years of smoking, and clinical center. We further adjusted spirometry traits for height and HRCT traits for scanner model. Longitudinal traits were also adjusted for their corresponding baseline trait values.

### Combined Multi-Omic Scores

For continuous traits, we constructed combined multi-omic risk scores (MultRS) using the multiview R package.^23^ We trained least absolute shrinkage and selection (LASSO) late fusion models and compared model performance to ORS using the aforementioned framework.

*P*-values were adjusted for multiple testing in each analysis using the Benjamini-Hochberg method for controlling false discovery rate, and an adjusted *p-*value ≤ 0·05 was considered statistically significant.^24^ All statistical analyses and data visualization were performed using R 4·4·2.^25^

## RESULTS

### Cohort Characteristics

We selected a study cohort of 3,399 COPDGene participants with complete clinical data and plasma metabolomics, proteomics, and whole blood transcriptomics collected at the second visit (which occurred approximately five years after baseline recruitment). After randomly splitting them (80:20) into training (n = 2,684) and held-out testing (n = 715) sets for model development and evaluation, respectively, they were used to develop and test MetRS, ProtRS, and TransRS for a range of COPD-related traits (**Supplementary Figure 1**). Demographics of the testing and training splits were similar (**Supplementary Table 1**), showing only a statistically, though not clinically, significant difference for pack-years of smoking (44·33 ± 24·44 vs. 46·59 ± 29·03, p = 0·004).

ORS were additionally tested for replication in two external cohorts with multi-omics data, SPIROMICS and MESA (**Supplementary Figure 1 & Supplementary Table 1**). Pairwise comparisons between cohort demographics are shown in **Supplemental Tables S3-S5**. Demographics of SPIROMICS and COPDGene were similar in terms of sex (p = 0·18) and smoking status (p = 0·73), but differed significantly in age, race, and GOLD stages (p < 0·001). As anticipated due to its design, MESA differed significantly from the two COPD cohorts in every demographic measure (**Supplementary Table 1**). In particular, MESA had large numbers of non-smokers (p < 0·001) and fewer pack-years of smoking (p < 0·001).

### ORS Predict Traits on Which They are Trained

We trained elastic net penalized regression models for each ORS, testing a range of values for α (mixing parameter that controls mixture between ℓ_1_ and ℓ_2_ penalization) to minimize mean-squared error. By this approach, α = 1 indicates a pure LASSO model and α = 0 indicates a pure ridge model (**Supplementary Figure 2A**). For 96% of ORS, LASSO models (which use purely ℓ_1_ penalization) performed within one standard error (of the MSE estimate) of the best performing elastic net model (**Supplementary Figure 2B**). We selected models within one standard error of the best performing model with maximum α and λ, resulting in sparser models with fewer features (**Supplementary Figure 1**). Resulting models contained between one and 7,708 features (**Supplementary Figure 2C**). The median number of features for final models was 104·5. Model parameters are displayed in **Supplementary Table S6**.

We evaluated the relationship between ORS and traits in the held-out COPDGene testing set using generalized linear models adjusting for age, sex, race, smoking status, pack-years of smoking, clinical center, plus height for spirometry traits and scanner model for HRCT traits. Traits of interest were significantly associated with 69 of 72 ORS (24 of 24 ProtRS, 23 of 24 MetRS, and 22 of 24 TransRS), with MetRS and TransRS for chronic bronchitis and TransRS for presence of exacerbations (both binary traits) not showing statistical significance (**Figure 1A** and **Supplementary Table 7**). For given traits, TransRS generally displayed smaller effect sizes (ANOVA *p =* 5·3 x 10^-5^) and were less significant (ANOVA *p =* 3·6 x 10^-4^ than MetRS and ProtRS).

**Figure 1.**
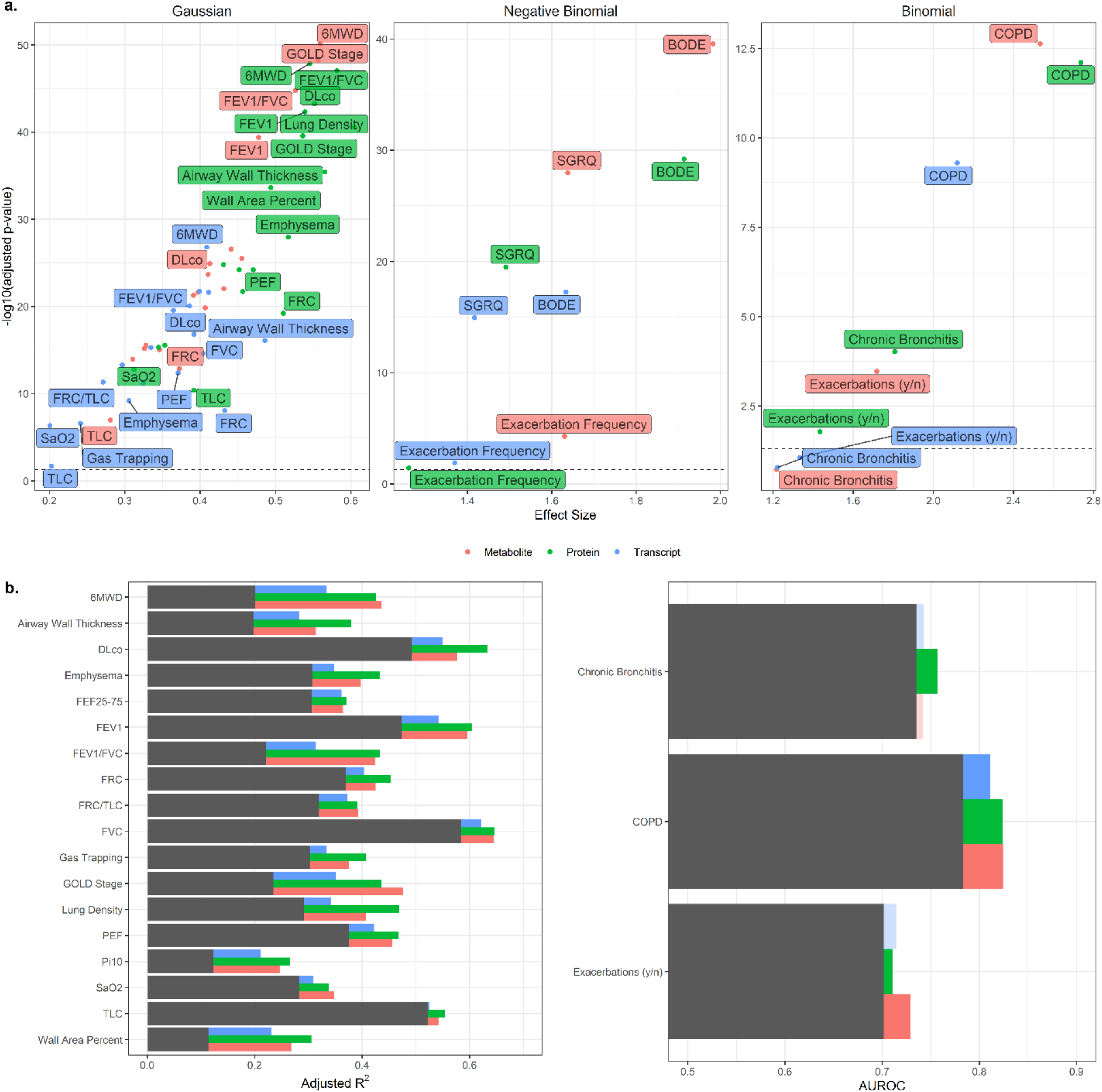
Performance of ORS in the COPDGene testing cohort. A) Score significance plotted against effect size for ORS associations with traits collected concurrently. Traits modeled with a Gaussian distribution are plotted with standardized β coefficient as the effect size measure while non-Gaussian-model traits are plotted with odds ratio. B) Adjusted R^2^ of trait models with and without ORS for traits measured concurrently with omics. Grey bars indicate variance explained by models including only clinical risk factors. AUROC of trait models with and without ORS.

### ORS Add Predictive Value to Clinical Models of COPD Traits

To determine the added value of ORS in trait prediction, we constructed a series of null models using demographic and clinical covariates in the COPDGene testing group (minimally age, sex, race, smoking status, pack years of smoking, and clinical center) and evaluated the performance of ORS when added to these models. By receiver-operator curve (ROC) analysis, significantly associated scores at baseline improved the area under the curve by an average increase of 0·022 (MetRS, 0·025; ProtRS, 0·024; TransRS, 0·016) (**Figure 1B**).

### ProtRS, MetRS, & TransRS for FEV_1_/FVC Predict COPD

Because COPD is currently defined by an abnormal FEV_1_/FVC ≤ 0·7, we focused on scores for that trait (**Figure 2**). ORS were significantly higher in COPD cases in the COPDGene testing group (adjusted *p*-value = 2·34 x 10^-13^, 7·87 x 10^-13^, 5·03 x 10^-10^ for MetRS, ProtRS, and TransRS, respectively) relative to controls (**Figure 2A**). ORS provided an increase in AUROC relative to a null model only considering clinical covariates (age, sex, race, clinical center, smoking status, and pack-years of smoking) of 0·83 for MetRS or ProtRS, and 0·81 for TransRS vs. 0·78 for the null model (**Figure 2B**). MetRS, ProtRS, and TransRS for COPD significantly added to model prediction (Delong’s p = 9·1 x 10^-6^, 4·6 x 10^-5^, and 8·5 x 10^-4^, respectively). Top features for each FEV_1_/FVC ORS are shown in **Figure 2C**.

**Figure 2.**
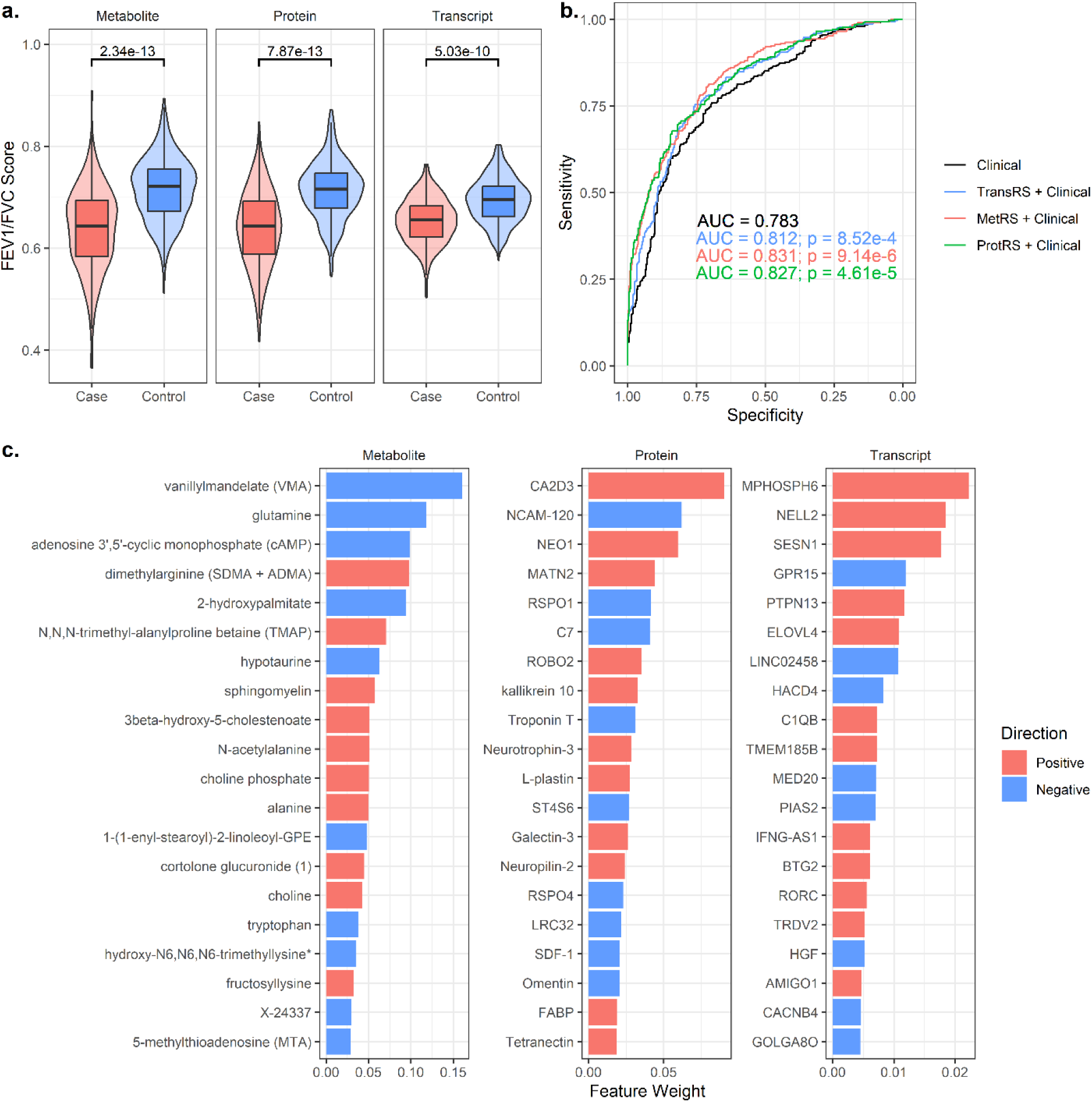
ORS improve prediction of COPD status. A) Violin plots of the FEV_1_/FVC MetRS, ProtRS, and TransRS stratified by COPD cases and controls. B) ROC plot of COPD predictive models including clinical covariates with or without the MetRS, ProtRS, or TransRS. C) Top 20 features in the MetRS, ProtRS, and TransRS, ranked by feature importance. Bars are colored by direction of association.

### ORS Replicate in External Cohorts

We next tested replication of the ORS derived in COPDGene against those traits available in the external study cohorts. SPIROMICS, had 14 overlapping traits (6MWD, emphysema, FEF_25-75_, FEV_1_, FEV_1_/FVC, FVC, GOLD stage, lung density, PEF, Pi10, resting oxygenation, TLC, chronic bronchitis, and COPD). For each of the 14 traits, all 38 ORS showed significant cross-sectional associations (**Figure 3A, Supplementary Table S8)**. For traits with linked transcriptomics data, significance estimates and variance explained for TransRS were lower than for MetRS and ProtRS (**Figure 3B-E**).

**Figure 3.**
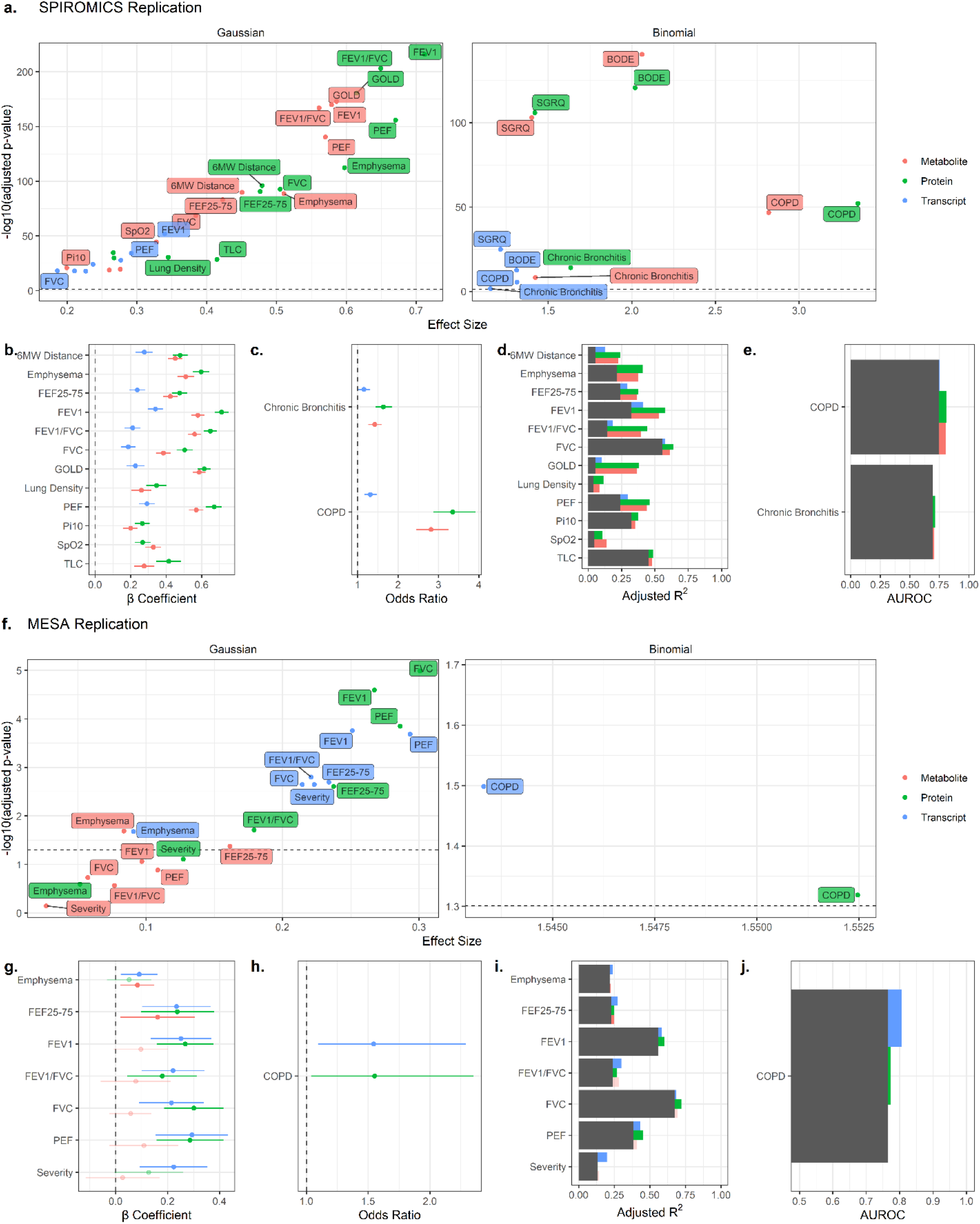
Replication of ORS in SPIROMICS & MESA. A) Score significance plotted against effect size for ORS tested in SPIROMICS. B) Standardized β coefficients and 95% confidence intervals for Gaussian trait model ORS in SPIROMICS. C) Odds ratios for binomial trait model ORS in SPIROMICS. D) Adjusted R^2^ for Gaussian trait model ORS in SPIROMICS. E) AUROC for binomial trait model ORS in SPIROMICS. F) Score significance plotted against effect size for ORS tested in MESA. G) Standardized β coefficients and 95% confidence intervals for Gaussian trait model ORS in MESA. H) Odds ratios for binomial trait ORS in MESA. I) Adjusted R^2^ for Gaussian trait model ORS in MESA. J) AUROC for binomial trait model ORS in MESA.

MESA had eight traits that overlapped with COPDGene (emphysema, FEF_25-75_, FEV_1_, FEV_1_/FVC, FVC, PEF, COPD, plus a severity score to which we applied our GOLD ORS). 16 out of 24 ORS were significant in MESA (two of eight MetRS, six of eight ProtRS, seven of eight TransRS) (**Figure 3F-J, Supplementary Table S9**). Only two MetRS replicated (for emphysema and FEF_25-75_), seemingly driven by the different analytic metabolomic platforms used between COPDGene and MESA, which shared few features. In contrast to the lesser performance of TransRS in the COPDGene testing set and SPIROMICS, in MESA TransRS showed similar effect sizes as ProtRS. This observation also likely resulted from limited feature overlap in ProtRS due to use of a smaller SomaScan proteomic platform in MESA.

Due to this concern, we went back and reconstructed ProtRS and MetRS for all traits in COPDGene, using only features present in the MESA data. Evaluating these new scores, we find that 7/8 ProtRS and all MetRS show significant association in MESA (**Supplementary Figure 4**).

### ORS Capture Trait Progression & Mortality

We next constructed ORS focused on disease progression, by training scores on the difference between Visits 2 and 3 of COPDGene in a subset of continuous traits (changes in 6MWD, FEV_1_

% predicted, total FEV_1_, annualized FEV_1_, and lung density) plus mortality. In contrast to ORS trained on cross-sectional traits, longitudinal scores were rarely associated with the trait of interest in the COPDGene testing set; only one of 15 ORS were significant (TransRS for change in FEV_1_ % predicted; **Figure 4A-B**; **Supplementary Table S10**).

**Figure 4.**
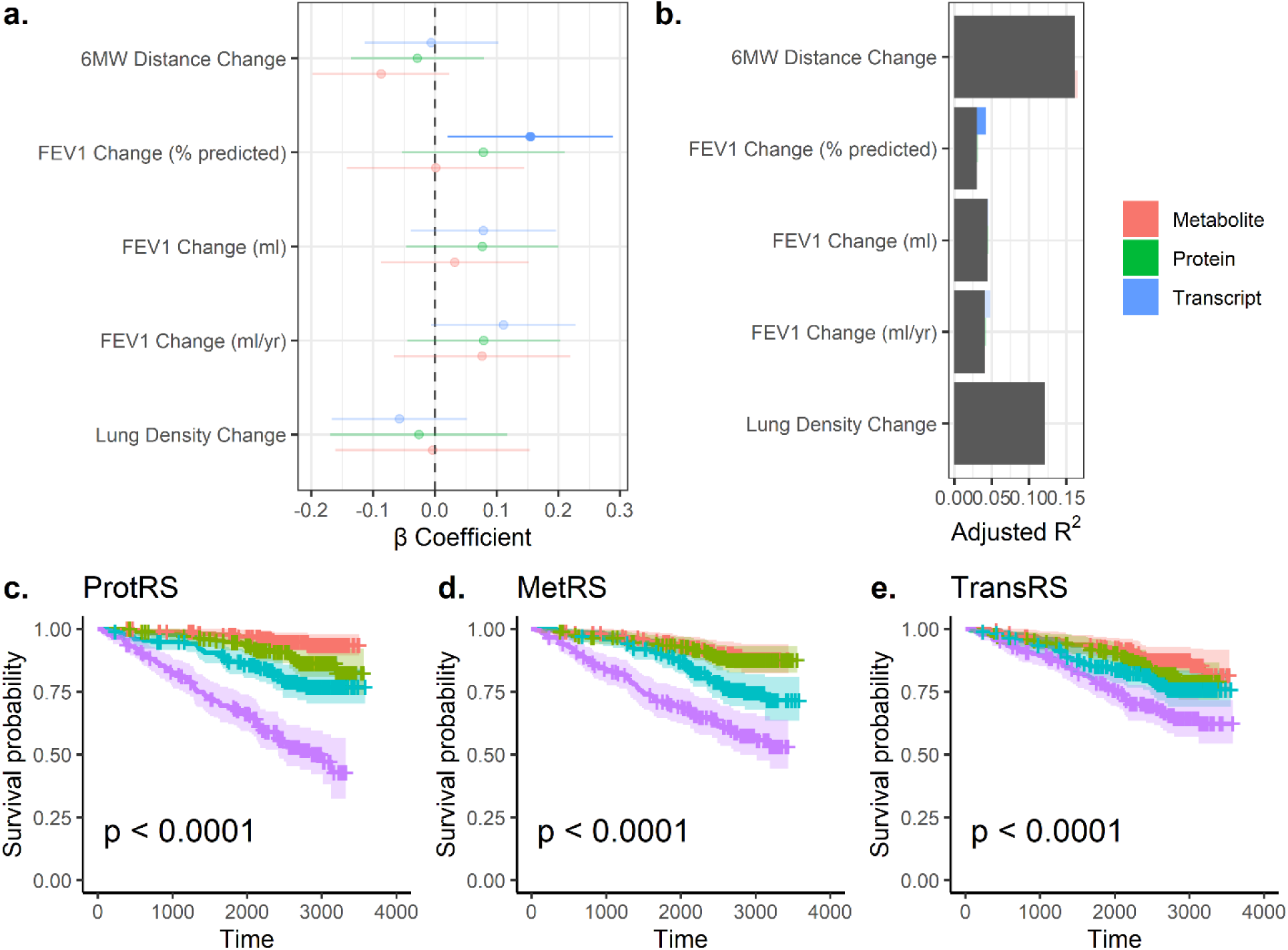
ORS of Progression Traits and Mortality in COPDGene. A) Standardized β coefficients and 95% confidence intervals for ORS. Darker bars indicate significant associations. B) Adjusted R^2^ for significant ORS in addition to clinical risk factors. C) Kaplan-Maier survival curve colored by ProtRS quartiles. D) Kaplan-Maier survival curve colored by MetRS quartiles. E) Kaplan-Maier survival curve colored by TransRS quartiles.

Mortality ORS were significantly associated with all-cause mortality in the held-out COPDGene testing set, with quartiles of all mortality ORS significantly associated with all-cause mortality (**Figure 4C-E**). Addition of both the MetRS and ProtRS individually to mortality models (which included age, sex, race, clinical center, smoking status, pack years of smoking, and BODE scores as covariates) significantly increased model performance (ProtRS, *p* = 2·04e-10; MetRS, 8·30e-4; TransRS, 0·056).

### Combined Multi-Omic Risk Scores for Continuous Traits

For continuous traits in the COPDGene testing set, we constructed combined multi-omic risk scores (MultRS) using late fusion to integrate features from all three omics data types into a single score (**Figure 5**; **Supplementary Table 11**). These combined multi-omic models consistently outperformed single-omic models (ANOVA *p* for difference in score βs = 1·4 x 10^-8^, ANOVA *p* for difference in score *p*-values = 2·3 x 10^-4^) (**Figure 5A-B**). Examining feature weights, many of the top-ranked features in single-omic scores were retained in the MultRS for lung density (**Figure 5C-F**). Notably, the top 60 features in MultRS encompassed nearly all top-ranking MetRS (20/20) and ProtRS (19/20) features but only three of the top 20 TransRS features

**Figure 5.**
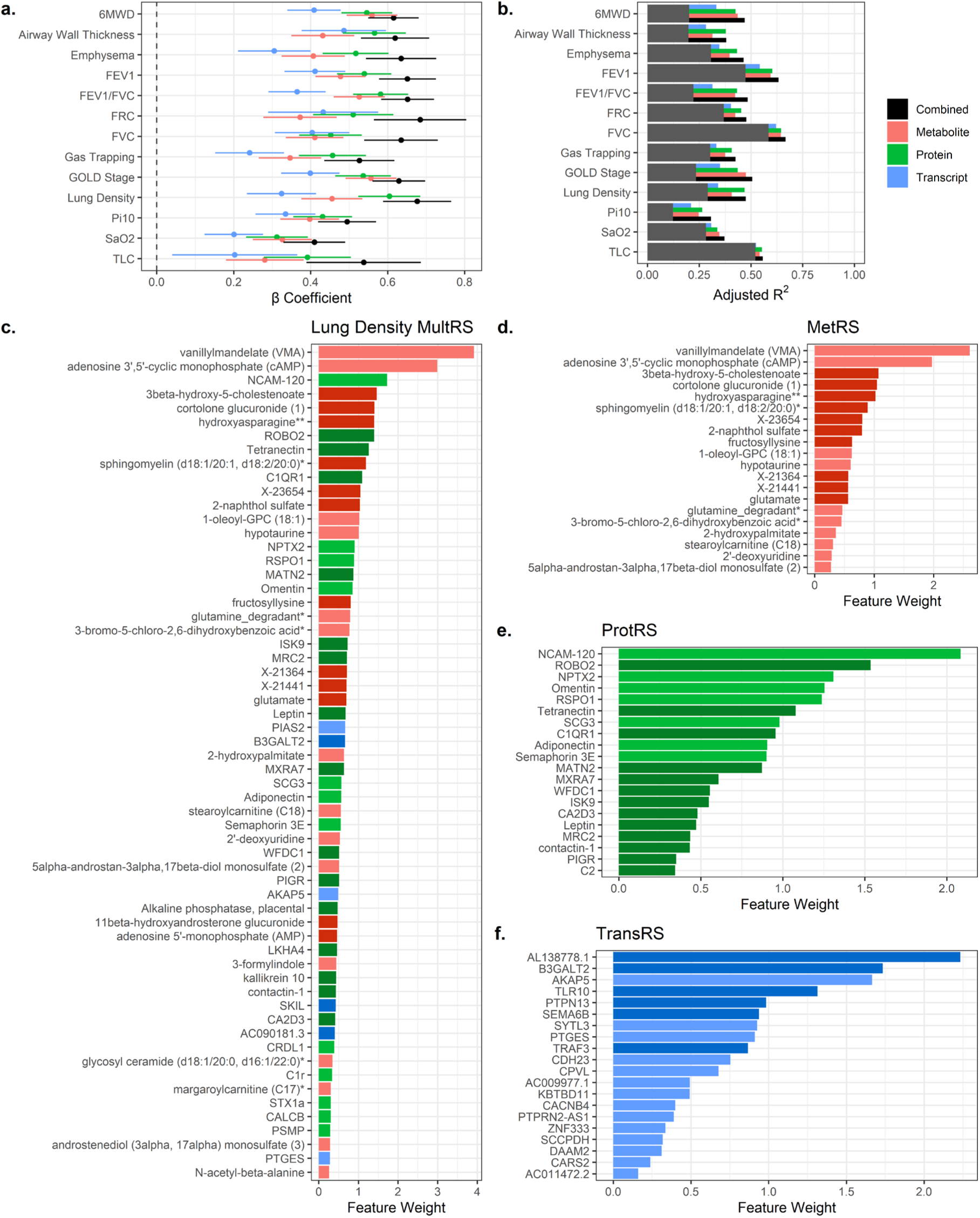
MultRS of Continuous COPDGene Traits. A) Standardized β coefficients and 95% confidence intervals for ORS and MultRS. B) Adjusted R^2^ for MultRS and ORS in addition to clinical risk factors. C) Top 60 features in the lung density MultRS. Positive associations are darkly shaded. D) Top 20 features in the lung density MetRS. E) Top 20 features in the lung density ProtRS. F) Top 20 features in the lung density TransRS.

## DISCUSSION

Using phenotypic and multi-omics data from thee large observational NHLBI cohorts, we constructed ORS, both single modality (MetRS, ProtRS, and TransRS) and multi-omic (MultRS), and analyzed them for cross-sectional and longitudinal associations with clinically relevant COPD outcomes. There are four principal findings. First, independently of demographic covariation, ORS were significantly associated with many cross-sectional traits indicating COPD disease severity in a variety of domains (physiological, imaging, symptoms). As indicated by the ROC analysis, ORS improved prediction of COPD status when added to clinical variables.

Second, replication of ORS was high, across both independent cohorts and analytical platforms; replication was complete for all ORS tested in SPIROMICS and for the majority in MESA. Third, additional ORS that we constructed in the COPDGene training set prospectively predicted longitudinal changes in FEV_1_, the current gold-standard of COPD progression, and all-cause mortality. Fourth, proteomics and metabolomics outperformed transcriptomics, and driven by those two methodologies, combined MultRS for continuous COPD traits outperformed single-omic ORS. These results have important implications for dissecting COPD pathogenesis and may aid in selecting potential participants to enroll in therapeutic trials of novel disease-modifying therapies.

Compared to PRS, ORS have several attractive features for inclusion in risk prediction models. Among them is the simplicity of discovery and validation. Unlike the thousands of participants typically required to develop PRS, ORS can be constructed using hundreds of individuals. PRS also remain stable throughout the lifetime, whereas ORS capture transient processes, potentially useful in disease subtyping and understanding shorter-term disease processes. Nevertheless, despite the capacity of omics signatures to change over time, we demonstrate here that ORS remain predictive of traits collected five years after omics measurements. Additionally, recent studies have shown that a non-trivial proportion of variation in omic measures is not explained by common genetic variation, indicating that omic features capture aspects of disease independent of genetic risk.^26^ Finally, ORS can potentially be leveraged to impute traits when unmeasured in additional populations.

Our ORS demonstrate feature overlap with previous ORS of COPD traits.^12, 13^ For example, top features in the Godbole et al. MetRS for FEV_1_ (such as vanillylmandelate) and the Moll et al. TransRS for FEV_1_/FVC (such as the top feature *GPR15*) were seen in our FEV_1_/FVC and lung density ORS. Importantly, despite ORS being optimized for trait prediction (i.e., not to identify feature-level associations with traits independent of relevant covariation), top features within our scores often reflect known trait biology. For example, among the top FEV_1_/FVC MetRS metabolites are molecules such as cAMP, which is targeted by current COPD therapeutics^27^ and sphingomyelin, which has previously been associated with COPD and emphysema.^28^ Replication of fewer ORS in MESA compared to SPIROMICS and the COPDGene testing set appears largely driven by differences in omics assays, resulting in limited feature overlap between studies. This issue was particularly pronounced for MetRS, where fewer than 20% of features were shared. To address this, we constructed ORS in COPDGene using only features shared with MESA, leading to strong replication, with only a single score failing to replicate. Additionally, weaker replication in MESA may stem from differences in cohort composition, as MESA is population-based, whereas COPDGene and SPIROMICS are enriched for heavy smoking histories.

Interestingly, relative to MetRS and ProtRS, TransRS displayed reduced effect sizes in the COPDGene testing set and SPIROMICS cohort. Differences in data processing and normalization across omics types and assays can contribute to variation in ORS performance. While lower relative effect sizes for TransRS might thus result from transcriptomic-specific data processing, we favor the interpretation that plasma-based metabolomics and proteomics provide greater power for modeling respiratory traits. Unlike proteomics and metabolomics, which measure circulating biomarkers in plasma, RNA-Seq captures intracellular transcripts. Reflecting this, we have found that TransRS more strongly predict blood cell proportions than MetRS and ProtRS (data not shown). Nonetheless, transcriptomics play a crucial role: RNA-Seq is an untargeted approach that captures all expressed transcripts at sufficient depth, whereas SomaScan proteomics assays and annotated metabolomics datasets are limited to predefined subsets of known features.

In contrast to the strong association between ORS and cross-sectional traits, many of the ORS we built on longitudinal change in trait values within the COPDGene discovery cohort failed to show significant association with the trait of interest. These results are unsurprising as disease progression during the roughly five years between visits was modest, with most participants not changing GOLD stage. The increased variability of using the difference in trait value between two timepoints likely also contributes. Our results suggest that ORS generated on single-timepoint omics data may have limited ability to predict modest changes in disease traits, especially when change in trait over time is limited.

Limitations to this work should be highlighted. We did not investigate or address participant heterogeneity. For many COPD-related traits, there are likely relevant subpopulations that should be considered. Furthermore, ORS are potentially confounded by factors such as genetic similarity, environmental exposures, and social determinants of health. If ORS are to be deployed broadly and across different populations, the impact of these factors must be assessed. Clinical translation of ORS needs to involve in-depth, careful evaluation of the optimal ORS for a given trait and address clinical implementation issues such as timing of measurement and cost-benefit ratios, amongst many other issues.

To conclude, we trained MetRS, TransRS, and ProtRS on a variety of COPD-related, HRCT, and spirometric traits in the COPDGene study, and most of these ORS were associated with the relevant trait in three validation groups (internally in a held-out testing set of COPDGene and externally in SPIROMICS and MESA). Our findings demonstrate the utility of blood-based omics to predict respiratory-related traits.

## Data and Code Availability

COPDGene proteomic and metabolomic data generated through the TOPMed program can be accessed through dbGaP accession phs000951·v5·p5 with an approved TOPMed proposal. COPDGene transcriptomic data can be accessed through an ancillary study request. MESA proteomic, transcriptomic, and metabolomic data is available through dbGaP accession phs001416·v3·p1. SPIROMICS RNA-Sequencing data generated through the TOPMed program are available through dbGaP accession phs001927·v1·p1 with an approved TOPMed proposal. SPIROMICS proteomic and metabolomic data can be accessed with an approved ancillary study. Feature weights for final omic risk scores as well as R scripts used to process data, analyze ORS, perform statistical analysis, and generate figures in this manuscript can be found at https://github.com/konigsbergi/OmicRiskScoresCOPD. Example scripts for generating ORS models are available as well.

## Acknowledgments

Molecular data for the Trans-Omics in Precision Medicine (TOPMed) program was supported by the National Heart, Lung and Blood Institute (NHLBI). RNASeq for “NHLBI TOPMed: SubPopulations and InteRmediate Outcome Measures In COPD Study (SPIROMICS)” (phs001927·v1·p1) was performed at the Northwest Genomics Center (HHSN268201600032I). Metabolomics for “NHLBI TOPMed: MESA and MESA Family AA-CAC” (phs001416·v3·p1) was performed at the Broad Institute and Beth Israel Metabolomics Platform (HHSN268201600038I). RNASeq for “NHLBI TOPMed: MESA and MESA Family AA-CAC” (phs001416·v3·p1) was performed at the Broad Institute Genomics Platform (HHSN268201600034I) and the Northwest Genomics Center (HHSN268201600032I). Core support including centralized genomic read mapping and genotype calling, along with variant quality metrics and filtering were provided by the TOPMed Informatics Research Center (3R01HL-117626-02S1; contract HHSN268201800002I). Core support including phenotype harmonization, data management, sample-identity QC, and general program coordination were provided by the TOPMed Data Coordinating Center (R01HL-120393; U01HL-120393; contract HHSN268201800001I). We gratefully acknowledge the studies and participants who provided biological samples and data for TOPMed. The COPDGene study (NCT00608764) is supported by NHLBI grants U01 HL089897 and U01 HL089856 and by NIH contract 75N92023D00011. COPDGene has also been supported by the COPD Foundation through contributions made to an Industry Advisory Committee that has included AstraZeneca, Bayer Pharmaceuticals, Boehringer-Ingelheim, Genentech, GlaxoSmithKline, Novartis, Pfizer, and Sunovion. The authors thank the SPIROMICS participants and participating physicians, investigators, study coordinators, and staff for making this research possible. More information about the study and how to access SPIROMICS data is available at www.spiromics.org. The authors would like to acknowledge the University of North Carolina at Chapel Hill BioSpecimen Processing Facility (http://bsp.web.unc.edu/) and Alexis Lab (https://www.med.unc.edu/cemalb/facultyresearch/alexislab/) for sample processing, storage, and sample disbursements. We would like to acknowledge the following current and former investigators of the SPIROMICS sites and reading centers: Neil E Alexis, MD; Wayne H Anderson, PhD; Mehrdad Arjomandi, MD; Igor Barjaktarevic, MD, PhD; R Graham Barr, MD, DrPH; Patricia Basta, PhD; Lori A Bateman, MS; Christina Bellinger, MD; Surya P Bhatt, MD; Eugene R Bleecker, MD; Richard C Boucher, MD; Russell P Bowler, MD, PhD; Russell G Buhr, MD, PhD; Stephanie A Christenson, MD; Alejandro P Comellas, MD; Christopher B Cooper, MD, PhD; David J Couper, PhD; Gerard J Criner, MD; Ronald G Crystal, MD; Jeffrey L Curtis, MD; Claire M Doerschuk, MD; Mark T Dransfield, MD; M Bradley Drummond, MD; Christine M Freeman, PhD; Craig Galban, PhD; Katherine Gershner, DO; MeiLan K Han, MD, MS; Nadia N Hansel, MD, MPH; Annette T Hastie, PhD; Eric A Hoffman, PhD; Yvonne J Huang, MD; Robert J Kaner, MD; Richard E Kanner, MD; Mehmet Kesimer, PhD; Eric C Kleerup, MD; Jerry A Krishnan, MD, PhD; Wassim W Labaki, MD; Lisa M LaVange, PhD; Stephen C Lazarus, MD; Fernando J Martinez, MD, MS; Merry-Lynn McDonald, PhD; Deborah A Meyers, PhD; Wendy C Moore, MD; John D Newell Jr, MD; Elizabeth C Oelsner, MD, MPH; Jill Ohar, MD; Wanda K O’Neal, PhD; Victor E Ortega, MD, PhD; Robert Paine, III, MD; Laura Paulin, MD, MHS; Stephen P Peters, MD, PhD; Cheryl Pirozzi, MD; Nirupama Putcha, MD, MHS; Sanjeev Raman, MBBS, MD; Stephen I Rennard, MD; Donald P Tashkin, MD; J Michael Wells, MD; Robert A Wise, MD; and Prescott G Woodruff, MD, MPH. The project officers from the Lung Division of the National Heart, Lung, and Blood Institute were Lisa Postow, PhD, and Lisa Viviano, BSN; SPIROMICS was supported by contracts from the NIH/NHLBI (HHSN268200900013C, HHSN268200900014C, HHSN268200900015C, HHSN268200900016C, HHSN268200900017C, HHSN268200900018C, HHSN268200900019C, HHSN268200900020C), grants from the NIH/NHLBI (U01 HL137880, U24 HL141762, R01HL182622, and R01 HL144718), and supplemented by contributions made through the Foundation for the NIH and the COPD Foundation from Amgen; AstraZeneca/MedImmune; Bayer; Bellerophon Therapeutics; Boehringer-Ingelheim Pharmaceuticals, Inc.; Chiesi Farmaceutici S.p.A.; Forest Research Institute, Inc.; Genentech; GlaxoSmithKline; Grifols Therapeutics, Inc.; Ikaria, Inc.; MGC Diagnostics; Novartis Pharmaceuticals Corporation; Nycomed GmbH; Polarean; ProterixBio; Regeneron Pharmaceuticals, Inc.; Sanofi; Sunovion; Takeda Pharmaceutical Company; and Theravance Biopharma and Mylan/Viatris. Funding for MESA lung measures is provided by NHLBI R01HL077612 and R01HL093081. Phenotype harmonization, data management, sample-identity quality control (QC), and general study coordination were provided by the TOPMed Data Coordinating Center (3R01HL-120393-02S1) and TOPMed MESA Multi-Omics (HHSN2682015000031/HSN26800004). The MESA projects are conducted and supported by the National Heart, Lung, and Blood Institute (NHLBI) in collaboration with MESA investigators (accession number phs000209.v13.p3). Support for the MESA projects are conducted and supported by the NHLBI in collaboration with MESA investigators. Support for MESA is provided by contracts 75N92020D00001, HHSN268201500003I, N01-HC-95159, 75N92020D00005, N01-HC-95160, 75N92020D00002, N01-HC-95161, 75N92020D00003, N01-HC-95162, 75N92020D00006, N01-HC-95163, 75N92020D00004, N01-HC-95164, 75N92020D00007, N01-HC-95165, N01-HC-95166, N01-HC-95167, N01-HC-95168, N01-HC-95169, UL1-TR-000040, UL1-TR-001079, UL1-TR-001420, UL1TR001881, DK063491, and R01HL105756. A full list of participating MESA investigators and institutes can be found at http://www.mesa-nhlbi.org.

## Author contributions

Conceptualization, IRK, EML, LAL, and MRM; Methodology, IRK and MRM; Formal Analysis, IRK; interpretation, IRK, LBV, EML, LAL, and MRM; writing – original draft: IRK; writing – revision, IRK, EML, LAL, MRM, EKS, and MHC; writing – review, supervision, LAL & MRM. All authors contributed to critical revision of the manuscript and read and approved the final manuscript.

