## Supplement for "Omic Risk Scores are Associated with COPD-related Traits Across Three Cohorts"

### Supplementary Materials

#### Table of Contents


| Title | Page(s) | Information |
|---|---|---|
| Supplementary Materials | 2-3 | TOPMed acknowledgements & supplementary methods |
| Supplementary Figure S1 | 4 | Schematic of study workflow. |
| Supplementary Figure S2 | 5 | Elastic net performance in ORS construction. |
| Supplementary Figure S3 | 6 | Elastic net cross-validation performance for predicting FEV1 in the COPDGene training set using proteomics data. |
| Supplementary Table S1 | 7 | Demographic comparisons between COPDGene training and testing datasets, SPIROMICS, and MESA |
| Supplementary Table S2 | 8 | Demographic comparisons between COPDGene training and testing datasets. |
| Supplementary Table S3 | 9 | Demographic comparisons between COPDGene and SPIROMICS. |
| Supplementary Table S4 | 10 | Demographic comparisons between COPDGene and MESA. |
| Supplementary Table S5 | 11 | Demographic comparisons between SPIROMICS and MESA. |
| Supplementary Table S6 | 12-13 | Hyperparameter selection for each cross-sectional trait model. |
| Supplementary Table S7 | 14-15 | ORS performance in the COPDGene testing dataset. |
| Supplementary Table S8 | 16 | ORS performance in SPIROMICS. |
| Supplementary Table S9 | 17 | ORS performance in MESA. |
| Supplementary Table S10 | 18 | Longitudinal ORS performance in the COPDGene testing dataset. |
| Supplementary Table S11 | 19 | MultRS performance in the COPDGene testing dataset. |

***TOPMed Banner Authors***

Christine Albert, Alvaro Alonso, Peter Anderson, Kristin Ardlie, Dan Arking, Donna K Arnett, Allison Ashley-Koch, Stella Aslibekyan, Tim Assimes, Najib Ayas, John Barnard, Kathleen Barnes, Emily Barron-Casella, Gerald Beck, Larry Bielak, John Blangero, Nathan Blue, Donald W. Bowden, Russell Bowler, Deborah Brown, Esteban Burchard, Carlos Bustamante, Brian Cade, Jonathan Cardwell, Julie Carrier, April P. Carson, Juan P Casas Romero, James Casella, Daniel Chasman, Yii-Der Ida Chen, Mina Chung, Clary Clish, Suzy Comhair, Joanne Curran, Brian Custer, Dawood Darbar, Colleen Davis, Michelle Daya, Mariza de Andrade, Michael DeBaun, Shannon Dugan-Perez, Jon Peter Durda, Susan K. Dutcher, Patrick Ellinor, Serpil Erzurum, Tasha Fingerlin, Myriam Fornage, Weiniu Gan, Bruce Gelb, Mark Geraci, Soren Germer, Robert Gerszten, Richard Gibbs, Mark Gladwin, Sharon Graw, Kathryn J. Gray, Xiuqing Guo, Jeff Haessler, Patrick Hanly, Nicola L. Hawley, Jiang He, Susan Heckbert, Ryan Hernandez, Bertha Hidalgo, James Hixson, John Hokanson, Marguerite Ryan Irvin, Cashell Jaquish, Jill Johnsen, Andrew Johnson, Craig Johnson, Robert Kaplan, Sharon Kardia, Shannon Kelly, Eimear Kenny, Barbara Konkle, Charles Kooperberg, Jiwon Lee, Dan Levy, Xiaohui Li, Henry Lin, Ruth J.F. Loos, Steven Lubitz, Kathryn Lunetta, James Luo, Ulysses Magalang, Michael Mahaney, Rasika Mathias, Stephen McGarvey, Becky McNeil, Luisa Mestroni, Ginger Metcalf, Deborah A Meyers, Emmanuel Mignot, Julie Mikulla, Ryan L Minster, Braxton D. Mitchell, Courtney Montgomery, Donna Muzny, Girish Nadkarni, Sergei Nekhai, Deborah Nickerson, Kari North, Allan Pack, Nicholette Palmer, George Papanicolaou, Patricia Peyser, Wendy Post, Michael Preuss, Bruce Psaty, Pankaj Qasba, Zhaohui Qin, Laura Raffield, D.C. Rao, Susan Redline, Catherine Reeves, Alex Reiner, Nicolas Robine, Dan Roden, Jerome Rotter, Sarah Ruuska, Danish Saleheen, Vijay G. Sankaran, David Schwartz, Christine Seidman, Jonathan Seidman, Vivien Sheehan, Stephanie L. Sherman, Wayne Hui-Heng Sheu, M. Benjamin Shoemaker, Jennifer Smith, Josh Smith, Nicholas Smith, Tamar Sofer, Nona Sotoodehnia, Kent D. Taylor, Matthew Taylor, Marilyn Telen, Sarah Tishkoff, Russell Tracy, David Van Den Berg, Daniel E. Weeks, L. Keoki Williams, Scott Williams, Joseph Wu, Lisa Yanek, Yingze Zhang, Elad Ziv, Michael Zody

#### ***Supplementary Methods***

*Omics Generation and Processing*

*RNA-Sequencing in COPDGene*

Subjects with available samples from the second COPDGene study visit were selected for RNA-Seq. Total RNA was extracted from PAXgene Blood RNA tubes using the Qiagen PreAnalytiX PAXgene Blood mRNA Kit. Globin reduction and cDNA library preparation for total RNA was performed with the Illumina TruSeq Stranded Total RNA with Ribo-Zero Globin kit. 75bp paired-end reads were generated on the Illumina HiSeq2000 platform. Samples were sequenced to an average depth of 22 million read pairs. Trimmed reads were aligned to GRCH38 genome using STAR. Samples were included for subsequent analysis if they had >10 million reads, >80% of reads mapped, XIST and Y chromosome expression consistent with reported gender, <10% of R1 reads in the sense orientation, Pearson correlation >= 0.9 with other samples in the same library prep batch, and concordant genotype calls between RNA reads and DNA genotyping. Reads were extracted from STAR aligned bam files using bedtools and pseudo-aligned to transcripts annotated in GENCODE version 37 using Salmon. Gene-level counts were calculated from Salmon isoforms with tximeta, adjusting for effective gene lengths. Counts were further corrected for library size with upper quantile normalization and a library preparation batch effect was removed from the data. Low count genes with less or equal than 100 samples having a CPM greater than 1 were filtered out.

*Proteomics in COPDGene*

Plasma protein levels were measured using the SomaScan 5K (V4.0) platform. Protein levels were natural-log-transformed prior to analysis.

*Metabolomics in COPDGene*

Plasma samples were profiled using Metabolon (Durham, NC, USA) Global Metabolomics Platform. Metabolite values were batch normalized by dividing by the median metabolite value for each metabolite within a batch. Metabolites were excluded if > 20% of samples were missing values. For metabolites missing in <20% of samples, missing values were imputed with k-nearest neighbor imputation (kNN; k = 10) using the R package ‘impute’.

*RNA-Sequencing in SPIROMICS*

Blood RNA-Sequencing was performed as part of the TOPMed study. RNA was extracted from whole blood by the Northwest Genome Center.

*Proteomics in SPIROMICS*

Proteomic data were generated on plasma samples using the SomaScan 7K (V4.1) platform through SPIROMICS ancillary study #AS136. Protein levels were natural-log-transformed prior to analysis.

*Metabolomics in SPIROMICS*

Metabolomic data were generated on plasma samples using the Metabolon Global Metabolomics platform through SPIROMICS ancillary study #AS096. Metabolite levels were natural-log-transformed prior to analysis.

*MESA omics*

Proteomics, transcriptomics, and metabolomics data were generated as part of the TOPMed project. RNA-Sequencing of PBMCs was performed at the Northwest Genomics Center at the University of Washington and the Broad Institute of MIT and Harvard.

*RNA-Sequencing in MESA*

RNA-Sequencing was performed as part of the TOPMed project. RNA-seq was conducted by two NHLBI-sequencing laboratories (the Northwest Genomics Center at the University of Washington and the Broad Institute of MIT and Harvard) on MESA participants using harmonized protocols. RNA libraries were prepared from at least 250 ng RNA using the Illumina TruSeq™ Stranded mRNA Kit and sequenced using the Illumina HiSeq 4000 (Illumina, San Diego, CA) platform, for a target depth of $\geq$40 M 2 $\times$ 101 bp paired-end reads. Alignment was performed using the TOPMed RNA-seq pipeline. Briefly, reads were aligned to GRCh38 with STAR and collapsed to the gene-level using RNASEQC v2 and the GENCODE 34 reference. Comprehensive pipeline information is provided at https://github.com/broadinstitute/gtex-pipeline/blob/master/TOPMed_RNAseq_pipeline.md. Log-transformed transcripts per million were used in analysis.

*Proteomics in MESA*

Protein levels were measured using the SomaScan 1.3K platform as part of the TOPMed program. Protein levels were natural-log-transformed prior to analysis.

*Metabolomics in MESA*

Metabolomics quantification occurred as part of the TOPMed program using the Broad Institute and Beth Israel Metabolomics Platform. Metabolite levels were natural-log-transformed prior to analysis.

*MultRS*

Multi-omic scores were generated using early fusion, late fusion, and coordinated learning with the *multiview* package. We observed similar performance, and only present late fusion models in the main text. Briefly, we trained least absolute shrinkage and selection (LASSO) models across a range of fusion strategies (early fusion, in which omics datasets are concatenated prior to fitting a predictive model ($\rho = 0$), late fusion, in which a predictive model is fit to each omics dataset and then combined for final prediction ($\rho = 1$), and cooperative learning, which is a mixture of early and late fusion ($\rho = 0.5$)). As different fusion strategies yielded models with similar performance, we present only late fusion models.

**Supplementary Figure S1. Schematic of study workflow.**

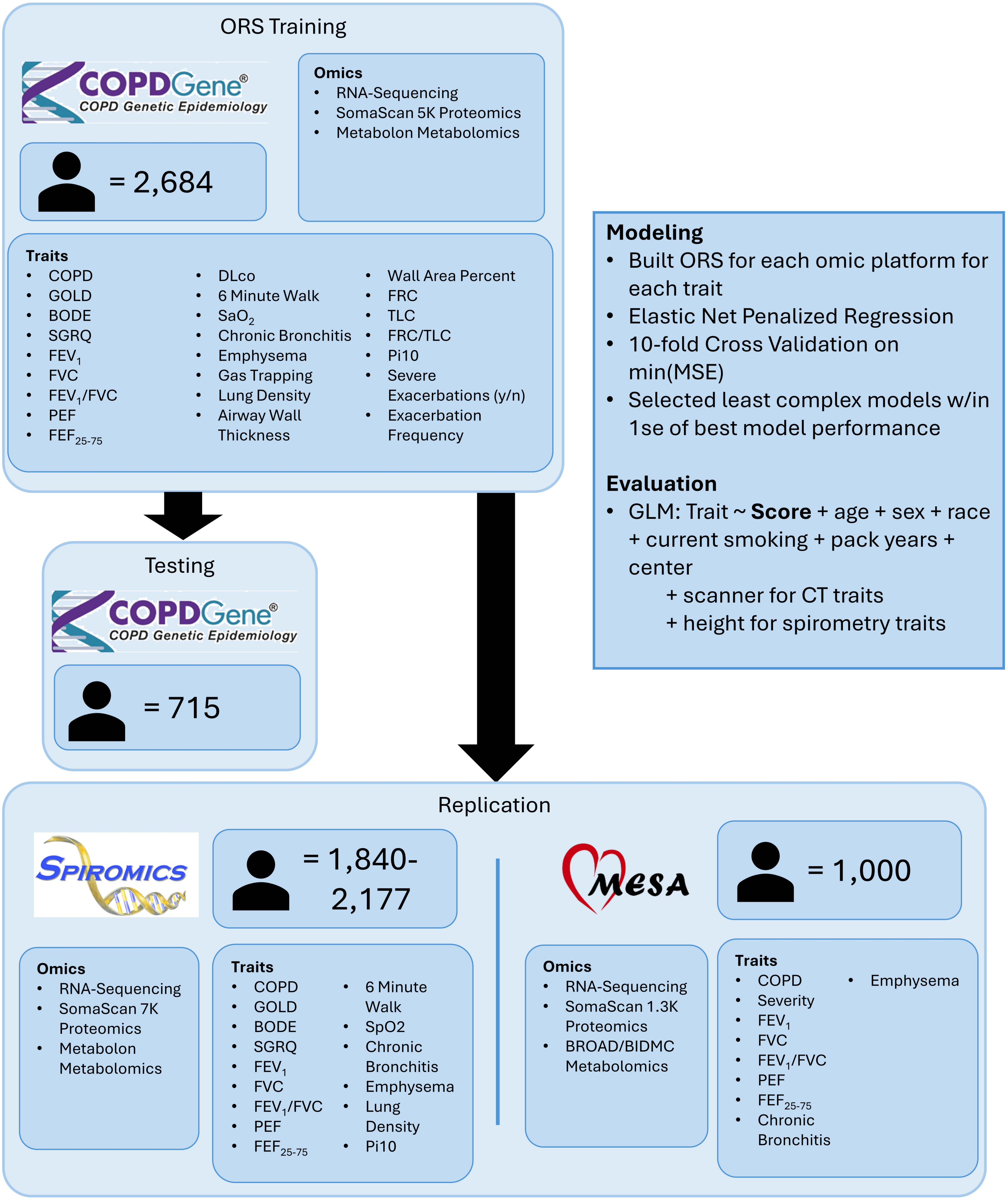

**Supplementary Figure S2. Elastic net performance in ORS construction. A) Number of features included in each final ORS. B) α (elastic net mixing parameter) selection for best-performing elastic net models. C) α selection for least-complex models with MSE within 1 standard error of best model performance.**

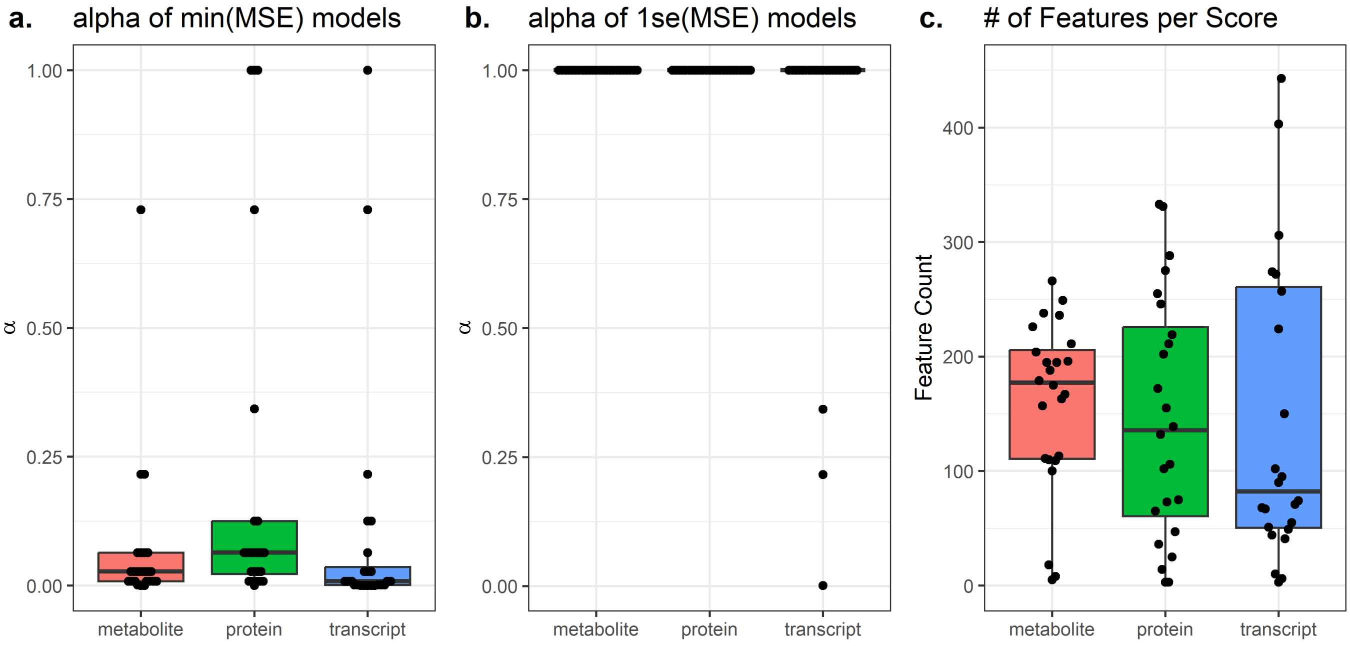

**Supplementary Figure S3. Elastic net cross-validation performance for predicting FEV1 in the COPDGene training set using proteomics data.**

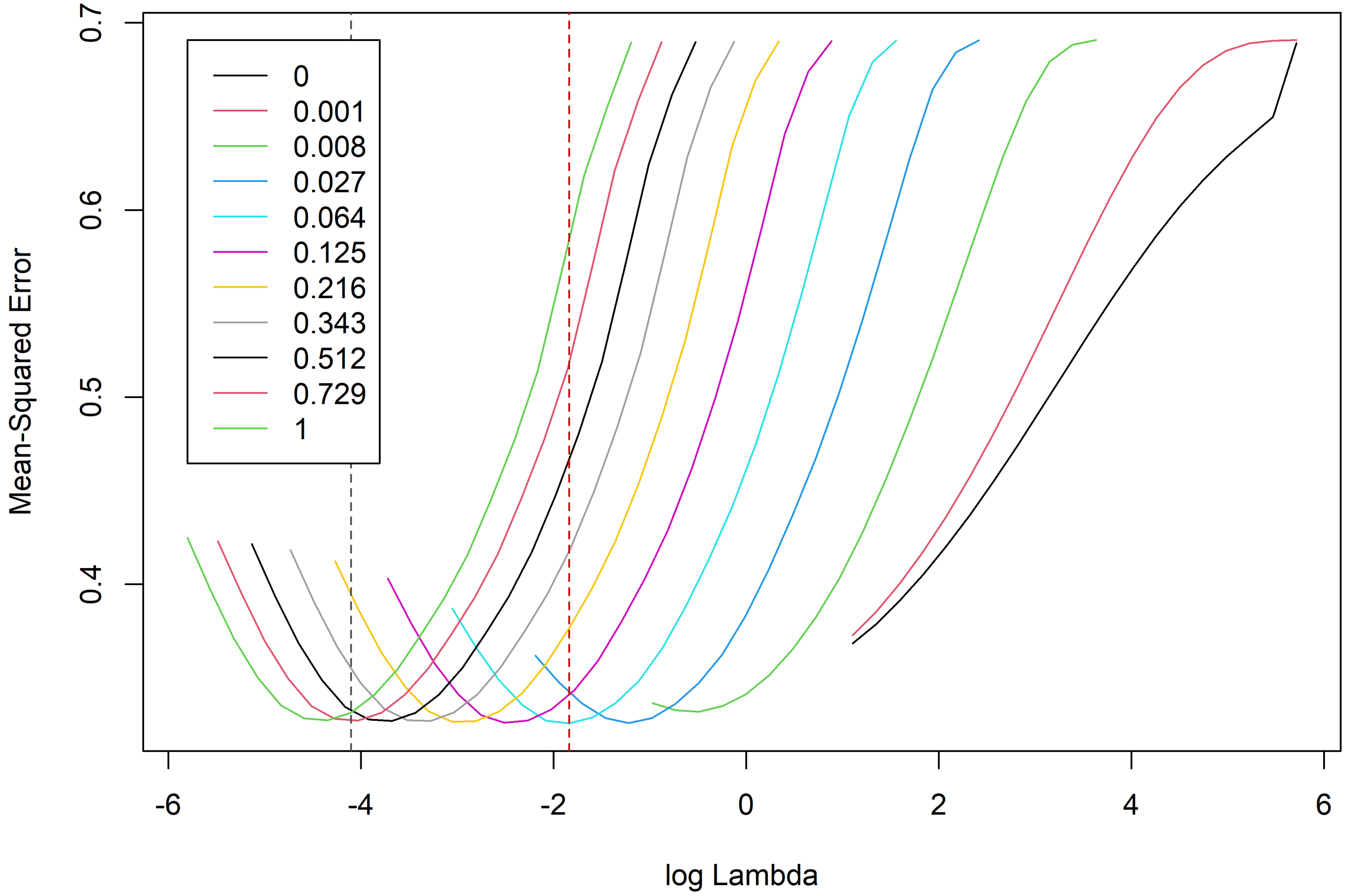

**Supplementary Table S1. Demographic comparisons between COPDGene training and testing datasets, SPIROMICS, and MESA**

| | **COPDGene Training** | **COPDGene Testing** | **SPIROMICS** | **MESA** | **p** |
|---|---|---|---|---|---|
| N | 2684 | 715 | 1659 | 1000 | |
| Age (mean (SD)) | 65.54 (8.74) | 65.50 (8.69) | 63.14 (9.14) | 69.70 (9.54) | <0.001 |
| Sex (No. (%) Male) | 1371 (51.1) | 374 (52.3) | 886 (53.4) | 468 (46.8) | 0.01 |
| Race (No. (%)) | | | | | <0.001 |
| American Indian or Alaska Native | 0 (0.0) | 0 (0.0) | 7 (0.4) | 0 (0.0) | |
| Asian | 0 (0.0) | 0 (0.0) | 13 (0.8) | 77 (7.7) | |
| Black or African American | 722 (26.9) | 186 (26.0) | 319 (19.3) | 196 (19.6) | |
| Hispanic | 0 (0.0) | 0 (0.0) | 0 (0.0) | 306 (30.6) | |
| Mixed | 0 (0.0) | 0 (0.0) | 41 (2.5) | 0 (0.0) | |
| White | 1962 (73.1) | 529 (74.0) | 1269 (77.0) | 421 (42.1) | |
| Smoking Status (No. (%)) | | | | | <0.001 |
| Current | 999 (37.2) | 261 (36.5) | 615 (37.6) | 78 (8.0) | |
| Former | 1685 (62.8) | 454 (63.5) | 1020 (62.4) | 412 (42.1) | |
| Never | 0 (0.0) | 0 (0.0) | 0 (0.0) | 488 (49.9) | |
| Pack-years of smoking (mean (SD)) | 43.80 (24.26) | 46.33 (24.99) | 46.59 (29.03) | 10.72 (18.95) | <0.001 |
| GOLD spirometry grades (No. (%)) | | | | | |
| Normal spirometry | 1491 (56.3) | 393 (55.4) | 509 (33.0) | | |
| 1 | 257 (9.7) | 84 (11.8) | 227 (14.7) | | |
| 2 | 520 (19.6) | 134 (18.9) | 461 (29.9) | | |
| 3 | 275 (10.4) | 68 (9.6) | 238 (15.4) | | |
| 4 | 107 (4.0) | 31 (4.4) | 107 (6.9) | | |

**Supplementary Table S2. Demographic comparisons between COPDGene training and testing datasets.**

| | COPDGene Testing | COPDGene Training | p |
|---|---|---|---|
| n | 715 | 2684 | |
| Age (mean (SD)) | 65.50 (8.69) | 65.54 (8.74) | 0.903 |
| Sex = Male (%) | 374 (52.3) | 1371 (51.1) | 0.588 |
| Race = White (%) | 529 (74.0) | 1962 (73.1) | 0.668 |
| Smoking Status = Former (%) | 454 (63.5) | 1685 (62.8) | 0.757 |
| Pack Years (mean (SD)) | 46.33 (24.99) | 43.80 (24.26) | 0.014 |
| GOLD (%) | | | 0.516 |
| Normal spirometry | 393 (55.4) | 1491 (56.3) | |
| 1 | 84 (11.8) | 257 ( 9.7) | |
| 2 | 134 (18.9) | 520 (19.6) | |
| 3 | 68 ( 9.6) | 275 (10.4) | |
| 4 | 31 ( 4.4) | 107 ( 4.0) | |

**Supplementary Table S3. Demographic comparisons between COPDGene and SPIROMICS.**

| | COPDGene | SPIROMICS | p |
|---|---|---|---|
| n | 3399 | 1659 | |
| Age (mean (SD)) | 65.54 (8.73) | 63.14 (9.14) | <0.001 |
| Sex = Male (%) | 1745 (51.3) | 886 (53.4) | 0.177 |
| Race (%) | | | <0.001 |
| American Indian or Alaska Native | 0 ( 0.0) | 7 ( 0.4) | |
| Asian | 0 ( 0.0) | 13 ( 0.8) | |
| Black or African American | 908 (26.7) | 319 (19.3) | |
| Mixed | 0 ( 0.0) | 41 ( 2.5) | |
| White | 2491 (73.3) | 1269 (77.0) | |
| Smoking Status = Former (%) | 2139 (62.9) | 1020 (62.4) | 0.731 |
| Pack Years (mean (SD)) | 44.33 (24.44) | 46.59 (29.03) | 0.004 |
| GOLD (%) | | | <0.001 |
| Normal spirometry | 1884 (56.1) | 509 (33.0) | |
| 1 | 341 (10.1) | 227 (14.7) | |
| 2 | 654 (19.5) | 461 (29.9) | |
| 3 | 343 (10.2) | 238 (15.4) | |
| 4 | 138 ( 4.1) | 107 ( 6.9) | |

**Supplementary Table S4. Demographic comparisons between COPDGene and MESA.**

| | COPDGene | MESA | p |
|---|---|---|---|
| n | 3399 | 1000 | |
| Age (mean (SD)) | 65.54 (8.73) | 69.70 (9.54) | <0.001 |
| Sex = Male (%) | 1745 (51.3) | 468 (46.8) | 0.013 |
| Race (%) | | | <0.001 |
| Asian | 0 ( 0.0) | 77 ( 7.7) | |
| Black or African America | n 908 (26.7) | 196 (19.6) | |
| Hispanic | 0 ( 0.0) | 306 (30.6) | |
| White | 2491 (73.3) | 421 (42.1) | |
| Smoking Status (%) | | | <0.001 |
| Current | 1260 (37.1) | 78 ( 8.0) | |
| Former | 2139 (62.9) | 412 (42.1) | |
| Never | 0 ( 0.0) | 488 (49.9) | |
| Pack Years (mean (SD)) | 44.33 (24.44) | 10.72 (18.95) | <0.001 |
| GOLD (%) | | | NaN |
| Normal spirometry | 1884 (56.1) | 0 ( NaN) | |
| 1 | 341 (10.1) | 0 ( NaN) | |
| 2 | 654 (19.5) | 0 ( NaN) | |
| 3 | 343 (10.2) | 0 ( NaN) | |
| 4 | 138 ( 4.1) | 0 ( NaN) | |

**Supplementary Table S5. Demographic comparisons between SPIROMICS and MESA.**

| | MESA | SPIROMICS | p |
|---|---|---|---|
| n | 1000 | 1659 | |
| Age (mean (SD)) | 69.70 (9.54) | 63.14 (9.14) | <0.001 |
| Sex = Male (%) | 468 (46.8) | 886 (53.4) | 0.001 |
| Race (%) | | | <0.001 |
| American Indian or Alaska Native | 0 ( 0.0) | 7 ( 0.4) | |
| Asian | 77 ( 7.7) | 13 ( 0.8) | |
| Black or African American | 196 (19.6) | 319 (19.3) | |
| Hispanic | 306 (30.6) | 0 ( 0.0) | |
| Mixed | 0 ( 0.0) | 41 ( 2.5) | |
| White | 421 (42.1) | 1269 (77.0) | |
| Smoking Status (%) | | | <0.001 |
| Current | 78 ( 8.0) | 615 (37.6) | |
| Former | 412 (42.1) | 1020 (62.4) | |
| Never | 488 (49.9) | 0 ( 0.0) | |
| Pack Years (mean (SD)) | 10.72 (18.95) | 46.59 (29.03) | <0.001 |
| GOLD (%) | | | NaN |
| Normal spirometry | 0 ( NaN) | 509 (33.0) | |
| 1 | 0 ( NaN) | 227 (14.7) | |
| 2 | 0 ( NaN) | 461 (29.9) | |
| 3 | 0 ( NaN) | 238 (15.4) | |
| 4 | 0 ( NaN) | 107 ( 6.9) | |

**Supplementary Table S6. Hyperparameter selection for each cross-sectional trait model.**

| | | 1se Models | | min(mse) Models | |
|---|---|---|---|---|---|
| **Trait** | **Ome** | **α** | **# Features** | **α** | **# Features** |
| Airway Wall Thickness | metabolite | 1 | 211 | 0 | 1014 |
| BODE | metabolite | 1 | 157 | 0.008 | 1014 |
| Chronic Bronchitis | metabolite | 1 | 8 | 0.008 | 1014 |
| COPD | metabolite | 1 | 249 | 0.027 | 1014 |
| 6MW Distance | metabolite | 1 | 113 | 0.001 | 1014 |
| DLCO | metabolite | 1 | 238 | 0.027 | 1014 |
| Exacerbation Frequency | metabolite | 1 | 5 | 0.064 | 1014 |
| FEF25-75 | metabolite | 1 | 204 | 0.008 | 1014 |
| FEV1/FVC | metabolite | 1 | 196 | 0.064 | 1014 |
| FEV1 | metabolite | 1 | 266 | 0.008 | 1014 |
| GOLD Stage | metabolite | 1 | 236 | 0.027 | 1014 |
| FRC | metabolite | 1 | 109 | 0.008 | 1014 |
| FRC/TLC | metabolite | 1 | 195 | 0.027 | 1014 |
| FVC | metabolite | 1 | 188 | 0.216 | 1014 |
| Lung Density | metabolite | 1 | 179 | 0.027 | 1014 |
| Emphysema | metabolite | 1 | 111 | 0.216 | 1014 |
| Gas Trapping | metabolite | 1 | 110 | 0.729 | 1014 |
| PEF | metabolite | 1 | 175 | 0.027 | 1014 |
| Pi10 | metabolite | 1 | 226 | 0.064 | 1014 |
| SaO2 | metabolite | 1 | 100 | 0.064 | 1014 |
| Severe Exacerbations | metabolite | 1 | 18 | 0.008 | 1014 |
| SGRQ | metabolite | 1 | 167 | 0.008 | 1014 |
| TLC | metabolite | 1 | 195 | 0.027 | 1014 |
| Wall Area Percent | metabolite | 1 | 163 | 0 | 1014 |
| Airway Wall Thickness | protein | 1 | 73 | 0.027 | 4979 |
| BODE | protein | 1 | 36 | 0.008 | 4979 |
| Chronic Bronchitis | protein | 1 | 14 | 1 | 4979 |
| COPD | protein | 1 | 172 | 0.064 | 4979 |
| 6MW Distance | protein | 1 | 65 | 0.008 | 4979 |
| DLCO | protein | 1 | 331 | 0.027 | 4979 |
| Exacerbation Frequency | protein | 1 | 3 | 0 | 4979 |
| FEF25-75 | protein | 1 | 75 | 0.008 | 4979 |
| FEV1/FVC | protein | 1 | 288 | 0.064 | 4979 |
| FEV1 | protein | 1 | 333 | 0.064 | 4979 |
| GOLD Stage | protein | 1 | 275 | 1 | 4979 |
| FRC | protein | 1 | 106 | 0.027 | 4979 |
| FRC/TLC | protein | 1 | 202 | 0.125 | 4979 |
| FVC | protein | 1 | 255 | 0.064 | 4979 |
| Lung Density | protein | 1 | 246 | 0.343 | 4979 |
| Emphysema | protein | 1 | 219 | 0.729 | 4979 |
| Gas Trapping | protein | 1 | 155 | 0.008 | 4979 |
| PEF | protein | 1 | 211 | 0.064 | 4979 |
| Pi10 | protein | 1 | 139 | 0.125 | 4979 |
| SaO2 | protein | 1 | 47 | 1 | 4979 |
| Severe Exacerbations | protein | 1 | 3 | 0.008 | 4979 |
| SGRQ | protein | 1 | 25 | 0.064 | 4979 |
| TLC | protein | 1 | 102 | 0.027 | 4979 |
| Wall Area Percent | protein | 1 | 132 | 0.064 | 4979 |
| Airway Wall Thickness | transcript | 1 | 49 | 0 | 19263 |
| BODE | transcript | 1 | 67 | 0.008 | 19263 |
| Chronic Bronchitis | transcript | 1 | 6 | 0.027 | 19263 |
| COPD | transcript | 1 | 150 | 0.001 | 19263 |
| 6MW Distance | transcript | 0.343 | 306 | 0.001 | 19263 |
| DLCO | transcript | 1 | 257 | 0.001 | 19263 |
| Exacerbation Frequency | transcript | 1 | 3 | 0.008 | 19263 |

| | | | | | |
|---|---|---|---|---|---|
| FEF25-75 | transcript | 1 | 224 | 0.001 | 19263 |
| FEV1/FVC | transcript | 1 | 95 | 1 | 19263 |
| FEV1 | transcript | 1 | 403 | 0.008 | 19263 |
| GOLD Stage | transcript | 1 | 272 | 0 | 19263 |
| FRC | transcript | 1 | 41 | 0.125 | 19263 |
| FRC/TLC | transcript | 0.001 | 8053 | 0 | 19263 |
| FVC | transcript | 1 | 274 | 0.216 | 19263 |
| Lung Density | transcript | 1 | 71 | 0.125 | 19263 |
| Emphysema | transcript | 1 | 90 | 0.729 | 19263 |
| Gas Trapping | transcript | 1 | 102 | 0.027 | 19263 |
| PEF | transcript | 0.216 | 443 | 0.008 | 19263 |
| Pi10 | transcript | 1 | 55 | 0.001 | 19263 |
| SaO2 | transcript | 1 | 51 | 0 | 19263 |
| Severe Exacerbations | transcript | 1 | 10 | 0.008 | 19263 |
| SGRQ | transcript | 1 | 44 | 0.027 | 19263 |
| TLC | transcript | 1 | 74 | 0.064 | 19263 |
| Wall Area Percent | transcript | 1 | 68 | 0 | 19263 |

**Supplementary Table S7. ORS performance in the COPDGene testing dataset.**

| trait | ome | pval | adj_pval | beta | beta_ci_hi | beta_ci_lo | rsq_partial | rsq_full | rsq_null | glm_family |
|---|---|---|---|---|---|---|---|---|---|---|
| 6MW Distance | Metabolite | 8.20E-53 | 4.18E-51 | 0.558889514 | 0.493335989 | 0.624443039 | 0.293860859 | 0.436111906 | 0.201449033 | gaussian |
| SaO2 | Metabolite | 3.26E-16 | 7.22E-16 | 0.326005503 | 0.249513401 | 0.402497605 | 0.0909365 | 0.347980465 | 0.282756886 | gaussian |
| GOLD Stage | Metabolite | 2.30E-51 | 5.88E-50 | 0.559486193 | 0.493532919 | 0.625439467 | 0.318793647 | 0.478786284 | 0.234866626 | gaussian |
| FEV1/FVC | Metabolite | 5.65E-47 | 9.61E-46 | 0.52721821 | 0.460708811 | 0.59372761 | 0.260734869 | 0.424370663 | 0.221349266 | gaussian |
| FEV1 | Metabolite | 3.20E-41 | 2.96E-40 | 0.477450212 | 0.412323735 | 0.54257669 | 0.231628399 | 0.595845938 | 0.474012234 | gaussian |
| FVC | Metabolite | 2.20E-25 | 9.34E-25 | 0.411251913 | 0.336781623 | 0.485722204 | 0.145605866 | 0.645361207 | 0.584923657 | gaussian |
| PEF | Metabolite | 1.40E-22 | 4.59E-22 | 0.391376515 | 0.315552632 | 0.467200398 | 0.12945019 | 0.456388382 | 0.375553688 | gaussian |
| DLco | Metabolite | 1.23E-26 | 5.72E-26 | 0.413157206 | 0.340783555 | 0.485530857 | 0.170648413 | 0.578281653 | 0.491508363 | gaussian |
| FEF25-75 | Metabolite | 2.89E-16 | 6.56E-16 | 0.321892781 | 0.246519907 | 0.397265654 | 0.092009613 | 0.369582905 | 0.305700694 | gaussian |
| Airway Wall Thickness | Metabolite | 2.71E-23 | 1.02E-22 | 0.426458519 | 0.345554011 | 0.507363028 | 0.144180997 | 0.313743505 | 0.198118736 | gaussian |
| Lung Density | Metabolite | 3.23E-39 | 2.74E-38 | 0.541551445 | 0.465793339 | 0.617309552 | 0.234207356 | 0.451800725 | 0.28413962 | gaussian |
| Emphysema | Metabolite | 7.69E-23 | 2.70E-22 | 0.423716993 | 0.342405546 | 0.50502844 | 0.141154372 | 0.404900233 | 0.307084658 | log |
| Gas Trapping | Metabolite | 1.70E-16 | 3.95E-16 | 0.350846145 | 0.269639252 | 0.432053039 | 0.10610762 | 0.37675098 | 0.302757675 | log |
| TLC | Metabolite | 1.25E-08 | 2.13E-08 | 0.294132269 | 0.194005818 | 0.39425872 | 0.048651965 | 0.544925427 | 0.521646836 | log |
| FRC | Metabolite | 2.71E-13 | 5.13E-13 | 0.364566382 | 0.268816067 | 0.460316698 | 0.084092487 | 0.422258587 | 0.369203535 | log |
| Pi10 | Metabolite | 1.23E-23 | 4.83E-23 | 0.398937449 | 0.323887667 | 0.473987231 | 0.146089328 | 0.251277568 | 0.12317301 | log |
| FRC/TLC | Metabolite | 9.63E-17 | 2.40E-16 | 0.329191591 | 0.253656853 | 0.404726329 | 0.107792862 | 0.392980718 | 0.31963155 | gaussian |
| Wall Area Percent | Metabolite | 3.41E-29 | 1.93E-28 | 0.447474352 | 0.373043052 | 0.521905653 | 0.180080617 | 0.273961156 | 0.114488457 | gaussian |
| Exacerbation Frequency | Metabolite | 1.89E-05 | 2.92E-05 | 1.644288472 | 1.309255693 | 2.065054667 | 0.024018993 | 1.89E-05 | 1.644288472 | negbinomial |
| SGRQ | Metabolite | 7.58E-31 | 4.55E-30 | 1.64943581 | 1.51236194 | 1.799404057 | -0.090931392 | 7.58E-31 | 1.64943581 | negbinomial |
| BODE | Metabolite | 1.88E-43 | 2.14E-42 | 2.009847612 | 1.809598257 | 2.238587615 | -0.00152439 | 1.88E-43 | 2.009847612 | negbinomial |
| Chronic Bronchitis | Metabolite | 0.108791277 | 0.133695304 | 1.244553361 | 0.950941247 | 1.625862918 | NA | 0.743700043 | 0.735142474 | binomial |
| Exacerbations (y/n) | Metabolite | 1.69E-07 | 2.73E-07 | 2.365071931 | 1.724373363 | 3.293553533 | NA | 0.765929714 | 0.701609265 | binomial |
| COPD | Metabolite | 8.03E-14 | 1.61E-13 | 2.534822209 | 1.998078393 | 3.257644759 | NA | 0.825022896 | 0.782978868 | binomial |
| 6MW Distance | Protein | 2.96E-51 | 6.04E-50 | 0.553338719 | 0.487236207 | 0.61944123 | 0.286287833 | 0.430064459 | 0.201449033 | gaussian |
| SaO2 | Protein | 2.67E-14 | 5.67E-14 | 0.316520231 | 0.236633362 | 0.396407101 | 0.079406809 | 0.339710874 | 0.282756886 | gaussian |
| GOLD Stage | Protein | 5.02E-42 | 5.12E-41 | 0.534646428 | 0.463336512 | 0.605956343 | 0.267500946 | 0.439540527 | 0.234866626 | gaussian |
| FEV1/FVC | Protein | 1.14E-51 | 3.86E-50 | 0.585681747 | 0.515942506 | 0.655420988 | 0.28369298 | 0.442247014 | 0.221349266 | gaussian |
| FEV1 | Protein | 1.26E-44 | 1.61E-43 | 0.538588564 | 0.468518122 | 0.608659005 | 0.248980537 | 0.60497295 | 0.474012234 | gaussian |
| FVC | Protein | 9.04E-26 | 4.01E-25 | 0.446672036 | 0.366489719 | 0.526854353 | 0.147806566 | 0.646274666 | 0.584923657 | gaussian |
| PEF | Protein | 7.38E-27 | 3.59E-26 | 0.475725417 | 0.392345913 | 0.559104922 | 0.153984973 | 0.471709037 | 0.375553688 | gaussian |
| DLco | Protein | 1.15E-45 | 1.68E-44 | 0.551537162 | 0.481458406 | 0.621615918 | 0.281903663 | 0.634854018 | 0.491508363 | gaussian |
| FEF25-75 | Protein | 2.10E-17 | 5.49E-17 | 0.360172462 | 0.279077449 | 0.441267475 | 0.098867406 | 0.374344266 | 0.305700694 | gaussian |
| Airway Wall Thickness | Protein | 4.45E-37 | 3.49E-36 | 0.565531606 | 0.483825944 | 0.647237267 | 0.225794238 | 0.379186801 | 0.198118736 | gaussian |
| Lung Density | Protein | 2.33E-58 | 2.38E-56 | 0.662645161 | 0.589919386 | 0.735370935 | 0.332380101 | 0.522078532 | 0.28413962 | gaussian |
| Emphysema | Protein | 1.44E-31 | 9.81E-31 | 0.522228914 | 0.439246206 | 0.605211622 | 0.19383288 | 0.441401517 | 0.307084658 | log |
| Gas Trapping | Protein | 5.47E-25 | 2.23E-24 | 0.470023938 | 0.38461245 | 0.555435427 | 0.162223488 | 0.415876673 | 0.302757675 | log |
| TLC | Protein | 1.95E-11 | 3.55E-11 | 0.392668342 | 0.279841768 | 0.505494916 | 0.06753344 | 0.553957326 | 0.521646836 | log |
| FRC | Protein | 1.13E-22 | 3.84E-22 | 0.53461741 | 0.431799415 | 0.637435404 | 0.147260777 | 0.462104244 | 0.369203535 | log |
| Pi10 | Protein | 6.53E-27 | 3.33E-26 | 0.43497544 | 0.359094609 | 0.510856271 | 0.166123198 | 0.268843584 | 0.12317301 | log |
| FRC/TLC | Protein | 1.60E-16 | 3.79E-16 | 0.345526114 | 0.26562949 | 0.425422739 | 0.106297656 | 0.391963445 | 0.31963155 | gaussian |
| Wall Area Percent | Protein | 2.33E-35 | 1.70E-34 | 0.494350776 | 0.42088855 | 0.567813002 | 0.21604425 | 0.305806964 | 0.114488457 | gaussian |
| Exacerbation Frequency | Protein | 5.86E-06 | 9.20E-06 | 1.623931841 | 1.31671158 | 2.002833926 | 0.017316576 | 5.86E-06 | 1.623931841 | negbinomial |
| SGRQ | Protein | 3.05E-20 | 8.88E-20 | 1.4842963 | 1.364354084 | 1.614851746 | -0.090931392 | 3.05E-20 | 1.4842963 | negbinomial |
| BODE | Protein | 6.08E-31 | 3.88E-30 | 1.917364575 | 1.712817618 | 2.151943114 | -0.00152439 | 6.08E-31 | 1.917364575 | negbinomial |
| Chronic Bronchitis | Protein | 5.96E-05 | 9.07E-05 | 1.807058287 | 1.358159172 | 2.422783896 | NA | 0.756696565 | 0.735142474 | binomial |
| Exacerbations (y/n) | Protein | 3.92E-06 | 6.24E-06 | 2.082401708 | 1.5364359 | 2.86834267 | NA | 0.749385873 | 0.701609265 | binomial |
| COPD | Protein | 1.05E-13 | 2.07E-13 | 2.768146897 | 2.1293254 | 3.645701188 | NA | 0.825387538 | 0.782978868 | binomial |
| 6MW Distance | Transcript | 3.40E-28 | 1.82E-27 | 0.407082547 | 0.337773414 | 0.476391679 | 0.164383265 | 0.332717448 | 0.201449033 | gaussian |
| SaO2 | Transcript | 7.30E-08 | 1.20E-07 | 0.210975194 | 0.13486846 | 0.287081928 | 0.039831336 | 0.311325638 | 0.282756886 | gaussian |
| GOLD Stage | Transcript | 4.34E-23 | 1.58E-22 | 0.397569363 | 0.321949812 | 0.473188914 | 0.151617447 | 0.350874195 | 0.234866626 | gaussian |
| FEV1/FVC | Transcript | 1.08E-21 | 3.34E-21 | 0.37147654 | 0.297822507 | 0.445130572 | 0.124270716 | 0.31811275 | 0.221349266 | gaussian |
| FEV1 | Transcript | 2.89E-22 | 9.22E-22 | 0.397104854 | 0.319542481 | 0.474667228 | 0.127608896 | 0.541132952 | 0.474012234 | gaussian |
| FVC | Transcript | 1.66E-15 | 3.59E-15 | 0.403743402 | 0.306551959 | 0.500934844 | 0.087423281 | 0.621210992 | 0.584923657 | gaussian |
| PEF | Transcript | 3.31E-14 | 6.88E-14 | 0.371114727 | 0.277109715 | 0.465119738 | 0.079510549 | 0.425203757 | 0.375553688 | gaussian |
| DLco | Transcript | 3.20E-19 | 9.07E-19 | 0.390716567 | 0.307954123 | 0.473479011 | 0.122958405 | 0.554031683 | 0.491508363 | gaussian |
| FEF25-75 | Transcript | 2.62E-13 | 5.04E-13 | 0.283969274 | 0.209240904 | 0.358697645 | 0.074009344 | 0.35708533 | 0.305700694 | gaussian |
| Airway Wall Thickness | Transcript | 1.47E-17 | 3.94E-17 | 0.480532485 | 0.373129564 | 0.587935406 | 0.107882273 | 0.284636608 | 0.198118736 | gaussian |
| Lung Density | Transcript | 6.03E-14 | 1.23E-13 | 0.368291611 | 0.274111787 | 0.462471435 | 0.082930008 | 0.343507529 | 0.28413962 | gaussian |
| Emphysema | Transcript | 1.49E-10 | 2.66E-10 | 0.309471982 | 0.216192544 | 0.40275142 | 0.061622865 | 0.34979233 | 0.307084658 | log |
| Gas Trapping | Transcript | 6.20E-08 | 1.04E-07 | 0.247510913 | 0.158844924 | 0.336176903 | 0.04632122 | 0.335066078 | 0.302757675 | log |
| TLC | Transcript | 0.009771245 | 0.014037564 | 0.210026176 | 0.050889772 | 0.369162581 | 0.00896628 | 0.525941895 | 0.521646836 | log |
| FRC | Transcript | 6.12E-09 | 1.06E-08 | 0.432713435 | 0.288658401 | 0.576768469 | 0.053484699 | 0.40295163 | 0.369203535 | log |
| Pi10 | Transcript | 5.57E-17 | 1.42E-16 | 0.34014489 | 0.262619832 | 0.417669948 | 0.103981234 | 0.214356523 | 0.12317301 | log |
| FRC/TLC | Transcript | 2.23E-12 | 4.14E-12 | 0.271841687 | 0.197376747 | 0.346306627 | 0.077718213 | 0.372519223 | 0.31963155 | gaussian |
| Wall Area Percent | Transcript | 2.16E-21 | 6.47E-21 | 0.38659711 | 0.309539986 | 0.463654234 | 0.132297545 | 0.231649233 | 0.114488457 | gaussian |
| Exacerbation Frequency | Transcript | 0.000170545 | 0.000255817 | 1.559158621 | 1.236853105 | 1.96545216 | 0.023982329 | 0.000170545 | 1.559158621 | negbinomial |
| SGRQ | Transcript | 1.12E-16 | 2.71E-16 | 1.430611633 | 1.308764999 | 1.564531637 | -0.090931392 | 1.12E-16 | 1.430611633 | negbinomial |
| BODE | Transcript | 1.56E-18 | 4.30E-18 | 1.636457215 | 1.462802595 | 1.835462354 | -0.00152439 | 1.56E-18 | 1.636457215 | negbinomial |

| | | | | | | | | | | |
|---|---|---|---|---|---|---|---|---|---|---|
| Chronic Bronchitis | Transcript | 0.05446943 | 0.071229255 | 1.291229113 | 0.993779756 | 1.675957394 | NA | 0.745820367 | 0.735142474 | binomial |
| Exacerbations (y/n) | Transcript | 0.003269075 | 0.004832546 | 1.567585307 | 1.164859755 | 2.12343634 | NA | 0.727502883 | 0.701609265 | binomial |
| COPD | Transcript | 3.33E-10 | 5.85E-10 | 2.093743576 | 1.670221336 | 2.650307041 | NA | 0.810589871 | 0.782978868 | binomial |

**Supplementary Table S8. ORS performance in SPIROMICS.**

| trait | ome | pval | adj_pval | beta | beta_ci_hi | beta_ci_lo | rsq_partial | rsq_full | rsq_null | glm_family |
|---|---|---|---|---|---|---|---|---|---|---|
| FEV1 | Protein | 1.03E-220 | 4.44E-219 | 0.712477974 | 0.67362537 | 0.751330578 | 0.382927753 | 0.321768012 | 0.581367753 | quant |
| FVC | Protein | 1.06E-96 | 3.04E-96 | 0.507576338 | 0.462351667 | 0.55280101 | 0.18585434 | 0.557024924 | 0.639255433 | quant |
| FEV1/FVC | Protein | 1.59E-213 | 3.42E-212 | 0.655706181 | 0.619190595 | 0.692221766 | 0.370850527 | 0.128967615 | 0.451841018 | quant |
| GOLD | Protein | 1.74E-190 | 2.50E-189 | 0.621298422 | 0.584438733 | 0.658158111 | 0.357783266 | 0.051811472 | 0.390888747 | quant |
| COPD | Protein | 1.60E-54 | 3.27E-54 | 3.362335149 | 2.893094694 | 3.928089147 | NA | 0.737902576 | 0.815703769 | binary |
| FEF25-75 | Protein | 8.42E-95 | 2.26E-94 | 0.482049953 | 0.438615648 | 0.525484258 | 0.180924107 | 0.242575133 | 0.379442401 | quant |
| PEF | Protein | 1.89E-162 | 1.16E-161 | 0.671471904 | 0.627220951 | 0.715722858 | 0.298958728 | 0.240610153 | 0.467491226 | quant |
| 6MW Distance | Protein | 1.34E-98 | 4.10E-98 | 0.483936506 | 0.441364412 | 0.526508599 | 0.198426406 | 0.055598406 | 0.242794821 | quant |
| Emphysema | Protein | 5.36E-120 | 2.10E-119 | 0.601585918 | 0.554798926 | 0.64837291 | 0.262366008 | 0.212792983 | 0.419019402 | quant |
| SpO2 | Protein | 2.26E-32 | 3.74E-32 | 0.27448309 | 0.229830968 | 0.319135213 | 0.067509006 | 0.046768869 | 0.11088859 | quant |
| TLC | Protein | 1.20E-29 | 1.79E-29 | 0.415859134 | 0.345047388 | 0.486670879 | 0.061464112 | 0.454152244 | 0.487429146 | quant |
| Lung Density | Protein | 1.47E-29 | 2.11E-29 | 0.326670229 | 0.270956183 | 0.382384276 | 0.073723871 | 0.039853896 | 0.110165396 | quant |
| Pi10 | Protein | 2.48E-34 | 4.26E-34 | 0.260843663 | 0.219865979 | 0.301821347 | 0.076181832 | 0.31986877 | 0.371347408 | quant |
| BODE | Protein | 1.11E-122 | 4.77E-122 | 2.022328646 | 1.898127921 | 2.15687034 | -0.00050025 | NA | NA | negbinom |
| SGRQ | Protein | 5.85E-106 | 1.94E-105 | 1.417829507 | 1.372474648 | 1.464809573 | -0.000535906 | NA | NA | negbinom |
| Chronic Broncl | Protein | 1.84E-15 | 2.03E-15 | 1.653326833 | 1.461836834 | 1.873254101 | NA | 0.693895118 | 0.71636224 | binary |
| FEV1 | Metabolite | 5.34E-173 | 4.59E-172 | 0.57998625 | 0.54311965 | 0.616852851 | 0.312502493 | 0.321768012 | 0.533637361 | quant |
| FVC | Metabolite | 1.45E-73 | 3.11E-73 | 0.389050903 | 0.348623395 | 0.429478411 | 0.137247289 | 0.557024924 | 0.617756615 | quant |
| FEV1/FVC | Metabolite | 1.22E-169 | 8.72E-169 | 0.562437772 | 0.526260537 | 0.598615008 | 0.305896525 | 0.128967615 | 0.395309875 | quant |
| GOLD | Metabolite | 1.53E-178 | 1.65E-177 | 0.590537865 | 0.553973093 | 0.627102637 | 0.336464417 | 0.051811472 | 0.370738698 | quant |
| COPD | Metabolite | 2.11E-48 | 3.94E-48 | 2.832692267 | 2.469271533 | 3.26485636 | NA | 0.737902576 | 0.805130813 | binary |
| FEF25-75 | Metabolite | 5.86E-92 | 1.48E-91 | 0.436109827 | 0.396095391 | 0.476124263 | 0.177484768 | 0.242575133 | 0.37689984 | quant |
| PEF | Metabolite | 1.89E-142 | 9.02E-142 | 0.5706486 | 0.529918298 | 0.611378901 | 0.264073526 | 0.240610153 | 0.441049219 | quant |
| 6MW Distance | Metabolite | 1.16E-89 | 2.62E-89 | 0.447525396 | 0.405936392 | 0.489114399 | 0.177948321 | 0.055598406 | 0.223531861 | quant |
| Emphysema | Metabolite | 1.82E-91 | 4.35E-91 | 0.511846083 | 0.465180698 | 0.558511467 | 0.207350505 | 0.212792983 | 0.375807432 | quant |
| SpO2 | Metabolite | 1.97E-45 | 3.52E-45 | 0.327609892 | 0.283290623 | 0.371929161 | 0.096866402 | 0.046768869 | 0.13897191 | quant |
| TLC | Metabolite | 9.78E-22 | 1.24E-21 | 0.283018865 | 0.225822895 | 0.340214836 | 0.05579945 | 0.450341891 | 0.480835276 | quant |
| Lung Density | Metabolite | 4.87E-18 | 5.51E-18 | 0.253912968 | 0.196993324 | 0.310832613 | 0.041422873 | 0.039853896 | 0.079311597 | quant |
| Pi10 | Metabolite | 6.28E-22 | 8.18E-22 | 0.199376569 | 0.159280186 | 0.239472952 | 0.043232592 | 0.324158711 | 0.353156259 | quant |
| BODE | Metabolite | 7.42E-149 | 3.99E-148 | 2.086535992 | 1.967733292 | 2.214459131 | -0.000489956 | NA | NA | negbinom |
| SGRQ | Metabolite | 8.24E-110 | 2.95E-109 | 1.409971215 | 1.365756377 | 1.455797981 | -0.000527426 | NA | NA | negbinom |
| Chronic Broncl | Metabolite | 2.81E-09 | 2.95E-09 | 1.434644638 | 1.274387842 | 1.617194879 | NA | 0.693895118 | 0.706817486 | binary |
| FEV1 | Transcript | 1.73E-53 | 3.37E-53 | 0.342345692 | 0.300328095 | 0.38436329 | 0.135866499 | 0.319621131 | 0.412061827 | quant |
| FVC | Transcript | 1.23E-18 | 1.47E-18 | 0.178364444 | 0.139145565 | 0.217583323 | 0.046313501 | 0.554454845 | 0.575089601 | quant |
| FEV1/FVC | Transcript | 7.79E-19 | 9.57E-19 | 0.21029717 | 0.164328579 | 0.256265761 | 0.046843485 | 0.14387734 | 0.183981109 | quant |
| GOLD | Transcript | 1.44E-18 | 1.68E-18 | 0.225099172 | 0.175540492 | 0.274657852 | 0.049407135 | 0.054642106 | 0.101349531 | quant |
| COPD | Transcript | 8.12E-06 | 8.32E-06 | 1.293561149 | 1.155835351 | 1.449357589 | NA | 0.745692471 | 0.751694954 | binary |
| FEF25-75 | Transcript | 3.94E-31 | 6.27E-31 | 0.267776884 | 0.223459845 | 0.312093923 | 0.079352922 | 0.243076494 | 0.303140585 | quant |
| PEF | Transcript | 8.82E-31 | 1.35E-30 | 0.283949107 | 0.236662072 | 0.331236142 | 0.078441482 | 0.225822325 | 0.286549969 | quant |
| 6MW Distance | Transcript | 2.11E-26 | 2.84E-26 | 0.262643227 | 0.215066333 | 0.31022012 | 0.068898121 | 0.054867238 | 0.119985109 | quant |
| BODE | Transcript | 1.75E-13 | 1.88E-13 | 1.311211656 | 1.221707134 | 1.408188562 | -0.000643087 | NA | NA | negbinom |
| SGRQ | Transcript | 1.38E-26 | 1.91E-26 | 1.219170282 | 1.175318064 | 1.264578461 | -0.000689655 | NA | NA | negbinom |
| Chronic Broncl | Transcript | 0.021320384 | 0.021320384 | 1.149453935 | 1.021148666 | 1.294661936 | NA | 0.693273818 | 0.694079665 | binary |

**Supplementary Table S9. ORS performance in MESA.**

| trait | ome | pval | adj_pval | beta | beta_ci_hi | beta_ci_lo | rsq_partial | rsq_full | rsq_null | glm_family |
|---|---|---|---|---|---|---|---|---|---|---|
| FEV1 | Protein | 3.52E-06 | 4.22E-05 | 0.260558476 | 0.152539365 | 0.368577588 | 0.100237074 | 0.55860971 | 0.599214227 | Gaussian |
| FVC | Protein | 2.30E-07 | 5.52E-06 | 0.300527256 | 0.189468364 | 0.411586148 | 0.153115071 | 0.676260258 | 0.7233174 | Gaussian |
| FEV1/FVC | Protein | 0.012667525 | 0.025335049 | 0.171680064 | 0.037051618 | 0.306308511 | 0.046678284 | 0.237290193 | 0.266229506 | Gaussian |
| Severity | Protein | 0.097857821 | 0.130477095 | 0.111575421 | -0.020689077 | 0.243839918 | 0.008149567 | 0.131696398 | 0.131769738 | Gaussian |
| Emphysema | Protein | 0.241674843 | 0.290009811 | 0.050226897 | -0.033908637 | 0.134362431 | -0.011931402 | 0.218224378 | 0.205588268 | Gaussian |
| COPD | Protein | 0.02294573 | 0.039335537 | 1.608010127 | 1.072949085 | 2.442877136 | NA | 0.765151515 | 0.777678571 | Binomial |
| FEF25-75 | Protein | 0.000989468 | 0.002638582 | 0.236903478 | 0.096980874 | 0.376826081 | 0.035125601 | 0.230202263 | 0.250418362 | Gaussian |
| PEF | Protein | 3.77E-05 | 0.000226419 | 0.273009026 | 0.144987393 | 0.40103066 | 0.110812751 | 0.384681106 | 0.447839923 | Gaussian |
| FEV1 | Metabolite | 0.054399353 | 0.081599029 | 0.10203203 | -0.001947108 | 0.206011167 | -0.048278705 | 0.55860971 | 0.527871901 | Gaussian |
| FVC | Metabolite | 0.294147949 | 0.320888672 | 0.043561022 | -0.038067525 | 0.125189568 | 0.051003485 | 0.676260258 | 0.689807776 | Gaussian |
| FEV1/FVC | Metabolite | 0.27707377 | 0.316655737 | 0.074018982 | -0.059962215 | 0.208000179 | 0.077839058 | 0.237290193 | 0.281748167 | Gaussian |
| Severity | Metabolite | 0.836180478 | 0.872536151 | 0.014927763 | -0.127337163 | 0.157192688 | 0.029869319 | 0.131696398 | 0.141290431 | Gaussian |
| Emphysema | Metabolite | 0.00826324 | 0.018028888 | 0.087079858 | 0.022517481 | 0.151642234 | 0.013876576 | 0.218224378 | 0.224679088 | Gaussian |
| COPD | Metabolite | 0.998015301 | 0.998015301 | NA | NA | NA | NA | NA | NA | Binomial |
| FEF25-75 | Metabolite | 0.07745215 | 0.109344212 | 0.125493974 | -0.013953892 | 0.26494184 | 0.036679784 | 0.230202263 | 0.242580392 | Gaussian |
| PEF | Metabolite | 0.120234794 | 0.15187553 | 0.105221584 | -0.02777984 | 0.238223008 | 0.057496078 | 0.384681106 | 0.407330262 | Gaussian |
| FEV1 | Transcript | 4.72E-05 | 0.000226419 | 0.240948979 | 0.126441576 | 0.355456381 | 0.059425618 | 0.55860971 | 0.58136001 | Gaussian |
| FVC | Transcript | 0.000734995 | 0.002204985 | 0.213250283 | 0.090434377 | 0.336066189 | 0.037528054 | 0.676260258 | 0.685798042 | Gaussian |
| FEV1/FVC | Transcript | 0.000434852 | 0.001739406 | 0.219149232 | 0.098148148 | 0.340150316 | 0.089160992 | 0.237290193 | 0.299471605 | Gaussian |
| Severity | Transcript | 0.000568522 | 0.001949217 | 0.230133748 | 0.100343614 | 0.359923883 | 0.086409365 | 0.131696398 | 0.20081955 | Gaussian |
| Emphysema | Transcript | 0.022023733 | 0.039335537 | 0.082054126 | 0.011847812 | 0.15226044 | 0.029565187 | 0.218224378 | 0.239158159 | Gaussian |
| COPD | Transcript | 0.024635431 | 0.03941669 | 1.52284457 | 1.083921656 | 2.276368406 | NA | 0.765151515 | 0.806287879 | Binomial |
| FEF25-75 | Transcript | 0.00120404 | 0.002889697 | 0.220937264 | 0.088128877 | 0.35374565 | 0.058489373 | 0.230202263 | 0.26913103 | Gaussian |
| PEF | Transcript | 1.92E-05 | 0.000153907 | 0.334804462 | 0.183565196 | 0.486043729 | 0.093533301 | 0.384681106 | 0.437542423 | Gaussian |

**Supplementary Table S10. Longitudinal ORS performance in the COPDGene testing dataset.**

| trait | ome | pval | adj_pval | beta | beta_ci_hi | beta_ci_lo | rsq_partial | rsq_full | rsq_null |
|---|---|---|---|---|---|---|---|---|---|
| 6MWD Change | Metabolite | 0.09592493 | 0.119321254 | -0.118619043 | -0.258443985 | 0.021205898 | 0.008823515 | 0.125377295 | 0.117591349 |
| FEV1 Change (ml) | Metabolite | 0.049946066 | 0.066162322 | 0.171532794 | 4.08E-05 | 0.343024829 | 0.013438436 | 0.050902536 | 0.037974417 |
| FEV1 Change (ml/yr) | Metabolite | 0.030666958 | 0.041158286 | 0.202640028 | 0.019064283 | 0.386215774 | 0.01731253 | 0.061717818 | 0.0451876 |
| FEV1 Change (% predicted) | Metabolite | 0.495045757 | 0.534386793 | 0.064189412 | -0.120940292 | 0.249319117 | -0.002519722 | 0.024636487 | 0.027087955 |
| Lung Density Change | Metabolite | 0.508383484 | 0.53493175 | -0.07788897 | -0.309834184 | 0.154056244 | -0.003225075 | 0.09112895 | 0.093992538 |
| 6MWD Change | Protein | 0.55420769 | 0.576828412 | -0.041127808 | -0.178009869 | 0.095754253 | -0.003223176 | 0.114747191 | 0.117591349 |
| FEV1 Change (ml) | Protein | 0.202895136 | 0.235173908 | 0.102854029 | -0.055879954 | 0.261588011 | 0.002970072 | 0.040831702 | 0.037974417 |
| FEV1 Change (ml/yr) | Protein | 0.169968855 | 0.201590967 | 0.110396636 | -0.047642831 | 0.268436102 | 0.004209337 | 0.049206728 | 0.0451876 |
| FEV1 Change (% predicted) | Protein | 0.014518979 | 0.020568554 | 0.187560436 | 0.037538095 | 0.337582777 | 0.023373782 | 0.049828589 | 0.027087955 |
| Lung Density Change | Protein | 0.694672811 | 0.708566267 | -0.037660073 | -0.226692841 | 0.151372695 | -0.004836748 | 0.089668856 | 0.093992538 |
| 6MWD Change | Transcript | 0.469204746 | 0.520205261 | 0.049348089 | -0.084840235 | 0.183536413 | -0.002352853 | 0.115515171 | 0.117591349 |
| FEV1 Change (ml) | Transcript | 0.016871892 | 0.023574424 | 0.208136505 | 0.037793338 | 0.378479672 | 0.022146945 | 0.059280344 | 0.037974417 |
| FEV1 Change (ml/yr) | Transcript | 0.025928543 | 0.035262818 | 0.169409616 | 0.020531058 | 0.318288173 | 0.01866199 | 0.0630063 | 0.0451876 |
| FEV1 Change (% predicted) | Transcript | 0.020164767 | 0.027794679 | 0.172478851 | 0.027241132 | 0.317716569 | 0.020696169 | 0.047223507 | 0.027087955 |
| Lung Density Change | Transcript | 0.857267712 | 0.865755511 | -0.013689864 | -0.163684274 | 0.136304545 | -0.005529986 | 0.089040818 | 0.093992538 |

**Supplementary Table S11. MultRS performance in the COPDGene testing dataset.**

| trait | ome | pval | adj_pval | beta | beta_ci_hi | beta_ci_lo | rsq_partial | rsq_full | rsq_null | glm_family |
|---|---|---|---|---|---|---|---|---|---|---|
| 6MWD | Combined | 5.48E-62 | 1.10E-60 | 0.615351358 | 0.550006036 | 0.680696681 | 0.336883199 | 0.470467437 | 0.201449033 | gaussian |
| FEV1 | Combined | 8.36E-56 | 4.18E-55 | 0.651223055 | 0.577251018 | 0.725195092 | 0.303310932 | 0.633550073 | 0.474012234 | gaussian |
| Airway Wall Thickness | Combined | 1.70E-37 | 3.10E-37 | 0.618977087 | 0.53014354 | 0.707810633 | 0.228144362 | 0.381071298 | 0.198118736 | gaussian |
| Lung Density | Combined | 9.14E-59 | 9.14E-58 | 0.728672275 | 0.649048334 | 0.808296215 | 0.33432643 | 0.523471828 | 0.28413962 | gaussian |
| Emphysema | Combined | 1.47E-37 | 2.93E-37 | 0.625563574 | 0.535868219 | 0.715258929 | 0.228193344 | 0.465210107 | 0.307084658 | log |
| 6MWD | Metabolite | 8.20E-53 | 3.28E-52 | 0.558889514 | 0.493335989 | 0.624443039 | 0.293860859 | 0.436111906 | 0.201449033 | gaussian |
| FEV1 | Metabolite | 3.20E-41 | 7.99E-41 | 0.477450212 | 0.412323735 | 0.54257669 | 0.231628399 | 0.595845938 | 0.474012234 | gaussian |
| Airway Wall Thickness | Metabolite | 2.71E-23 | 3.61E-23 | 0.426458519 | 0.345554011 | 0.507363028 | 0.144180997 | 0.313743505 | 0.198118736 | gaussian |
| Lung Density | Metabolite | 3.23E-39 | 7.17E-39 | 0.541551445 | 0.465793339 | 0.617309552 | 0.234207356 | 0.451800725 | 0.28413962 | gaussian |
| Emphysema | Metabolite | 7.69E-23 | 9.61E-23 | 0.423716993 | 0.342405546 | 0.50502844 | 0.141154372 | 0.404900233 | 0.307084658 | log |
| 6MWD | Protein | 2.96E-51 | 9.86E-51 | 0.553338719 | 0.487236207 | 0.61944123 | 0.286287833 | 0.430064459 | 0.201449033 | gaussian |
| FEV1 | Protein | 1.26E-44 | 3.61E-44 | 0.538588564 | 0.468518122 | 0.608659005 | 0.248980537 | 0.60497295 | 0.474012234 | gaussian |
| Airway Wall Thickness | Protein | 4.45E-37 | 7.42E-37 | 0.565531606 | 0.483825944 | 0.647237267 | 0.225794238 | 0.379186801 | 0.198118736 | gaussian |
| Lung Density | Protein | 2.33E-58 | 1.56E-57 | 0.662645161 | 0.589919386 | 0.735370935 | 0.332380101 | 0.522078532 | 0.28413962 | gaussian |
| Emphysema | Protein | 1.44E-31 | 2.22E-31 | 0.522228914 | 0.439246206 | 0.605211622 | 0.19383288 | 0.441401517 | 0.307084658 | log |
| 6MWD | Transcript | 3.40E-28 | 4.86E-28 | 0.407082547 | 0.337773414 | 0.476391679 | 0.164383265 | 0.332717448 | 0.201449033 | gaussian |
| FEV1 | Transcript | 2.89E-22 | 3.40E-22 | 0.397104854 | 0.319542481 | 0.474667228 | 0.127608896 | 0.541132952 | 0.474012234 | gaussian |
| Airway Wall Thickness | Transcript | 1.47E-17 | 1.63E-17 | 0.480532485 | 0.373129564 | 0.587935406 | 0.107882273 | 0.284636608 | 0.198118736 | gaussian |
| Lung Density | Transcript | 6.03E-14 | 6.34E-14 | 0.368291611 | 0.274111787 | 0.462471435 | 0.082930008 | 0.343507529 | 0.28413962 | gaussian |
| Emphysema | Transcript | 1.49E-10 | 1.49E-10 | 0.309471982 | 0.216192544 | 0.40275142 | 0.061622865 | 0.34979233 | 0.307084658 | log |
